# Correcting for volunteer bias in GWAS uncovers novel genetic variants and increases heritability estimates

**DOI:** 10.1101/2022.11.10.22282137

**Authors:** Sjoerd van Alten, Benjamin W. Domingue, Jessica Faul, Titus Galama, Andries T. Marees

**Affiliations:** Vrije Universiteit Amsterdam; Tinbergen Institute; Stanford University; University of Michigan; University of Southern California, Dornsife Center for Economic and Social Research and Department of Economics; Erasmus University Rotterdam, Erasmus School of Economics

**Keywords:** Genome wide association studies, selection bias, volunteer bias, participation bias, ascertainment bias, collider bias, polygenic scores, SNP-based heritability, gene-set analysis, tissue expression, MAGMA, LD-score regression, UK Biobank, inverse probability weights, genetic correlations

## Abstract

The implications of selection bias due to volunteering (volunteer bias) for genetic association studies are poorly understood. Because of its large sample size and extensive phenotyping, the UK Biobank (UKB) is included in almost all large genomewide association studies (GWAS) to date, as it is one of the largest cohorts. Yet, it is known to be highly selected. We develop inverse probability weighted GWAS (WGWAS) to estimate GWAS summary statistics in the UKB that are corrected for volunteer bias. WGWAS decreases the effective sample size substantially compared to GWAS by an average of 61% (from 337,543 to 130,684) depending on the phenotype. The extent to which volunteer bias affects GWAS associations and downstream results is phenotype-specific. Through WGWAS we find 11 novel genomewide significant loci for type 1 diabetes and 3 for breast cancer. These loci were not identified previously in any prior GWAS. Further, genetic variant’s effect sizes and heritability estimates become more predictive in WGWAS for certain phenotypes (e.g., educational attainment, drinks per week, breast cancer and type 1 diabetes). WGWAS also alters biological annotation relations in gene-set analyses. This suggests that not accounting for volunteer-based selection can result in GWASs that suffer from bias, which in turn may drive spurious associations. GWAS consortia may therefore wish to provide population weights for their data sets or rely more on population-representative samples.

## 1 Introduction

Genomewide association studies (GWAS) have resulted in the discovery of numerous genetic associations that can be used to facilitate our understanding of the genetic factors that contribute to variation in human phenotypes [1, 2]. However, as with other associations derived from non-representative data [3], GWAS results could be affected by selection bias, as individuals who volunteer to participate in a GWAS cohort are different from the underlying cohort-specific sampling population [3–10]. Under such circumstances, the internal validity of GWAS results may be affected, as study participation in itself can serve as a collider^1^ from genotype to phenotype. Here, we study whether this type of selection bias, which we will refer to as *volunteer bias*,^2^ affects GWAS findings for various phenotypes in the UK Biobank (UKB). Because the UKB is included in almost all large GWAS to date, as it is one of the largest cohorts with both rich genetic and rich phenotypic data, our findings are highly relevant to understanding biases in extant GWASs.

There are indications that genetic studies are affected by non-random selection. For example, sex shows significant autosomal heritability in data sets that require active participation (23andMe and the UKB), but not in data sets that require more passive enrollment [6]. As no known biological mechanism could cause autosomal allele frequencies to differ between the sexes, such observed autosomal heritability of sex can be attributed to sex-differential participation bias. Also, direct evidence exists that genes are associated with study engagement [12, 17]. For example, the UKB collects additional questionnaires and conducts follow-up studies that are optional. Selection into these optional components is significantly heritable [5, 7]. However, it is unclear whether (1) sample non-representativeness biases GWAS associations, and (2) whether and how non-representativeness also biases various downstream analyses based on such GWAS results as an input (e.g., SNP-based heritabilities or gene-set tissue expression analyses). We attempt to provide answers to such questions here.

To this end, we estimate GWAS results that are robust to volunteer bias and study the extent to which non-random selection into genotyped samples leads to bias. Theoretically, non-random sample selection may bias single nucleotide polymorphism (SNP) associations in various ways. Table 1 shows the results of simulations illustrating how selection may bias the regression of a phenotype *Y* on a hypothetical SNP of interest in various scenarios. *Phenotype-based selection* (scenario 1) results in attenuation bias and thus smaller SNP effect sizes. This results in potential false negatives, and may also affect various downstream analyses based on GWAS summary statistics, e.g., lead to smaller SNP heritabilities and alter bio-annotation results based on gene set analyses. *Phenotype-genotype-based selection* (scenario 2) is arguably more worrisome. Here, the sign of the bias depends on the sign by which the phenotype and SNP of interest *independently* influence selection into the sample. This type of selection can result in false positives when the true SNP effect size is zero, or can result in incorrect effect sizes (possibly of opposite sign) for SNPs that *do* have an effect on Y. When biases are sufficiently large, they may hamper our understanding of the genetic architecture of particular phenotypes.

**Table 1:** Simulated example of spurious SNP associations due to volunteer bias. We simulate a population of 1,000,000 individuals, with simulated phenotype *Y* = 0.04 · *SNP* + *ε*, with *SNP ∼* binom(1, 000, 000; 2; 0.4) and *ε∼ N* (0, 1). In this population, the true effect of the SNP on *Y* is 0.04. The effect of the SNP on *Y* is properly identified in the full population (see column 1; standard errors in parentheses). However, next we consider a non-randomly selected subsample that consists of 5% of this population, consisting of only those with 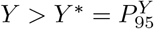 (*scenario 1: Phenotype-based selection*). Here, selection leads to attenuation bias in the SNP effect (column 2). Under an alternative selection scheme (*scenario 2: Phenotype-genotype-based selection*), selecting those with 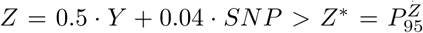, selection leads to downward bias so severe that the estimated SNP effect is of the wrong sign (column 3). In Column 4, the regression is estimated after selecting those with 0.5·*Y* − 0.04 · *SNP > Z*^*∗*^. As a result, the estimated SNP effect is upward biased.

|  | Phenotype |  |  |  |
| --- | --- | --- | --- | --- |
|  | Population | Scenario 1 | Scenario 2a | Scenario 2b |
|  | (1) | (2) | (3) | (4) |
| SNP | 0.041<br>(0.001) | 0.006<br>(0.002) | -0.062<br>(0.002) | 0.075<br>(0.002) |
| Constant | -0.00004<br>(0.002) | 2.088<br>(0.003) | 2.147<br>(0.003) | 2.033<br>(0.002) |
| Observations | 1,000,000 | 50,000 | 50,000 | 50,000 |
| R<sup>2</sup> | 0.001 | 0.0001 | 0.013 | 0.019 |

The UKB is a crucial data source for GWAS given its large sample size (*N* ≈ 500, 000) and extensive phenotyping. Due to the polygenicity of complex traits, typically small effect sizes of individual genetic variants and the large numbers of independent variants tested, large sample sizes are essential for GWASs to have sufficient power [18]. This makes the UKB one of the most important data sets in statistical genetics. However, the UKB suffers from selective participation: only 5.5% of UK citizens who received an invitation actually participated [13]. During data collection, the focus was on ensuring a large sample size, at the expense of ensuring representation of the underlying sampling population.^3^

In a related paper, we describe how this selective participation in the UKB results in substantial biases in various phenotype-phenotype associations, that can even be of the incorrect sign. Here, we use the inverse probability weights (IP weights) developed in that paper to correct estimated associations in the UKB for volunteer bias [3]. For the construction of such weights it is essential that the reference data source (i) is representative of the sampling population, (ii) has many variables in common with the target data source that relate to participation into the target data source (here the UKB), (iii) has sufficient sample size for reasons of precision, and (iv) has fine geographic detail. To our knowledge, only the UK Census (5% microdata subsample) meets all four criteria as a reference data source [19]. The UK Census is representative of the UK population (criterion i), has many variables in common with the UKB that relate to UKB participation (criterion ii) and has a substantial sample size (criterion iii). Last, the UKB restricted its sampling population to those living in sufficient proximity to any of 22 assessment centers, which were mainly located in highly urbanized areas. The fine geographic detail (criterion iv) of the UK Census data allowed us to precisely identify the sampling population of the UKB, by restricting to only individuals in the UKB-eligible age range that resided within the areas from which the UKB sampled its respondents. With these restrictions in place, this UKB-eligible subsample of the UK Census 5% microdata subsample (final sample size: 687,491) was used as an appropriate reference data source to estimate IP weights for all UKB respondents.^4^ As a result of meeting these criteria, these IP weights are precisely estimated and consequently capture an average of 78% of the volunteer bias in various estimated phenotype-phenotype associations.[3] Hence, by weighting the UKB using IP weights, we can estimate any association estimate as if the UKB were representative of its underlying sampling population.

Here, we use these IP weights to assess the extent to which GWAS results are biased due to non-representative sampling. We do this by running an inverse probability weighted GWAS (WGWAS) for 10 phenotypes. By comparing WGWAS and GWAS results, we provide evidence that non-random selection biases the estimated associations between genetic predictors and various phenotypes towards the null. In GWAS, volunteer bias results in missing genomewide significant SNP associations for type 1 diabetes and breast cancer, and in downward biased estimates of SNP-based heritabilities. For example, the SNP-based heritability of educational attainment (EA) in the UKB increases from 14.8 to 17.8 percent after accounting for volunteer bias. Further, contrasting WGWAS and GWAS results we find that volunteer bias can lead to misleading biological annotations.

These findings suggest that volunteer bias impacts genetic associations of interest as well as downstream uses of such estimated genetic effects. Because of the need for very large samples in GWAS, the focus has thus far been on growth in sample sizes, bringing together academic research samples from different countries, cohorts, and specific subpopulations, as well as samples collected by companies such as 23andMe. As a result, various types of selection biases may affect results based on such data sets. Our findings suggests that future GWASs may benefit from more systematic data collection that ensures representativeness (expensive) or from ensuring that sampling weights can be constructed to correct for potential biases (cheaper, and ideally incorporated as part of the study design for each included cohort).

To facilitate researchers in employing our volunteer bias correction, the IP weights will be made available to researchers using the UKB as a data field.

## 2 Results

### 2.1 WGWAS

To assess how selection into the UKB biases various phenotypes of interest, we performed a weighted GWAS (WGWAS) on 10 phenotypes. WGWAS estimates the association between the respective phenotype and the SNP through weighted least squares regression using IP weights designed to correct for volunteer bias (see Methods). These IP weights are available for ∼ 98% of UKB respondents (see [3]). We study the effects of selection into the UKB sample that is typically used in GWAS studies by restricting to individuals of British European Ancestry and removing those with low quality genetic data, or for whom IP weights could not be estimated (see Methods). For comparison, we estimate a corresponding GWAS for each phenotype, in which these regressions are left unweighted. Both WGWAS and GWAS control for genetic sex, the first 20 PCs of the genetic data, year of birth fixed effects, and gene batch fixed effects. Weighting introduces heteroskedasticity in standard errors. Therefore, we estimate heteroskedasticity robust standard errors for *both* WGWAS and GWAS.

Our final sample consists of 376,193 respondents. For reasons of computational feasibility, we restrict our analyses to 1,025,058 SNPs identified in HapMap3 that were available in the UKB imputed genotyped data set (call rate *>* 2%, MAF *>* 1% and in HWE with *p <* 1 · 10^*−*6^, as recommended in [21]). We selected 10 phenotypes related to health and social scientific outcomes. Supplementary table 1 summarizes these phenotypes before and after IP weighting. Weighting these phenotypes changes their mean and standard deviation.^5^ The sample size for all ten phenotypes is larger than 140,081, with an average N of 337,562 and maximal N of 376,900. Supplementary section S1 outlines our coding procedures for each phenotype.

To assess whether our IP weights capture volunteer-based selection that may affect phenotype-genotype associations, we first performed a GWAS with the IP weights as a phenotype. This resulted in 7 independent genomewide significant hits (Figure S1) and a SNP-based heritability of 3.6% (s.e. 0.26 %, LD-score intercept: 1.31 (0.0094)). The qq-plot for the associations shows an early lift-off (*λ* = 1.55; Figure S2). This implies that volunteer bias is highly polygenic and may impact genetic associations across the whole genome. Supplementary note S2 provides various additional analyses of our GWAS on the IP weights: the 7 top hits are found in loci for which significant genetic associations with various phenotypes have been reported, such as EA, cholesterol, alcohol consumption and hypothyroidism (supplementary note S2 and Figure S15). Further, these GWAS results exhibit substantial genetic overlap between the genes that influence a respondent’s IP weight and various other phenotypes, as measured by publicly available GWAS results (supplementary note S2 and Figure S3): genes associated with a higher IP weight are correlated with lower educational attainment (*rG* = −0.711 (0.0250)), a lower age at first birth (*rG* = −0.698 (0.0293)), a lower likelihood to participate in UKB’s optional modules (e.g. the mental health questionnaire, *rG* = −0.507 (0.0339)), and a higher BMI (*rG* = 0.265 (0.0226)), likelihood of smoking (*rG* = 0.395 (0.0267)) and likelihood of mental disorders (e.g., Depression *rG* = 0.288 (0.0333)). This pattern shows that the IP weights capture “healthy volunteer bias”, as they reflect that those in better health and of higher socioeconomic status (i.e., higher educational attainment) are more likely to volunteer for UKB participation (Figure S3).

Turning to the association results for phenotypes, we first investigate the relation between WGWAS and GWAS SNP effects for the (approximately independent) top hits for each phenotype, which we take from the literature. Here, we define a “top hit” as having *p <* 10^*−*5^ as determined by a well-powered GWAS taken from the literature (*N >* 200, 000) that did *not* include the UKB (see Methods for additional detail). Because well-powered GWAS that do not include UKB data are not available for every phenotype, we could only perform these analyses for 6 out of the 10 phenotypes. Using this set of SNPs, Table 2 shows the coefficient of a regression of the effect sizes estimated through WGWAS on the effect sizes estimated through GWAS. This coefficient is significantly larger than one for all cases, except for Breast Cancer. This implies that, for most phenotypes, correcting GWAS for volunteer bias through WGWAS results in *more predictive* effect sizes, i.e., effect sizes lie further from the null, which is consistent with selection bias, here taking the form of attenuation bias (i.e., selection is phenotype-based, scenario 1 from Table 1). Years of education, BMI, drinks per week, and severe obesity are most affected by this type of volunteer bias: correcting for volunteer bias results in an increase of the SNP effect sizes by 10.9% for years of education, 9.1% for BMI, 8.2% for severe obesity, and 18.3% for drinks per week. By contrast, correcting GWAS results for height for volunteer bias also results in significantly increased predictability, but the overall effect is relatively minor: a 2.1% increase in the effect sizes. This is intuitive, given that height only has a small genetic correlation with our IP weights (Figure S3), and it seems reasonable that height has little influence on whether individuals volunteer for scientific data participation.

**Table 2:** Comparison of weighted and unweighted GWAS results (top hits only). Each row shows the coefficient (and 95% confidence interval) for a bivariate regression with the weighted SNP effect as the dependent variable, and the unweighted SNP effect as the independent variable. A coefficient larger than one implies that WGWAS increases GWAS effect sizes on average (i.e., volunteer bias leads to an underestimate of the association in GWAS). A coefficient smaller than one implies that WGWAS shrinks effect sizes on average. P-values are for the null hypothesis that this coefficient equals one. The last column shows the number of SNPs that are included in this regressions: only independent top hits from GWAS studies that did *not* include the UKB are included (see Methods for additional detail).

| Phenotype | Coefficient [95% CI] | P | N |
| --- | --- | --- | --- |
| Years of Education | **1.109** [1.087;1.131] | $5.16 \times 10^{-21}$ | 504 |
| BMI | **1.091** [1.068;1.115] | $2.89 \times 10^{-13}$ | 259 |
| Severe Obesity | **1.082** [1.028;1.137] | 0.00300 | 259 |
| Height | **1.021** [1.014;1.028] | $3.83 \times 10^{-9}$ | 1967 |
| Drinks Per Week | **1.183** [1.054;1.312] | 0.00705 | 30 |
| Breast cancer | **0.794** [0.759;0.828] | $5.93 \times 10^{-28}$ | 510 |

Breast Cancer is the only phenotype for which we find a significant *shrinkage* of SNP effect sizes in Table 2, as the coefficient on the regression is 0.794. Hence, not taking volunteer bias into account inflates genetic effect sizes for previously identified top hits for breast cancer, which implies that some of these previously identified SNPs may have overestimated effect sizes. This is consistent with phenotype-genotype-based selection (scenario 2 in Table 1) for breast cancer. Out of the six phenotypes, breast cancer showed the largest change in prevalence after weighting: unweighted prevalence in the UKB was 2.9%, while weighted prevalence was 2.4%, a 16.2% change (supplementary table 1). Breast cancer could be particularly affected by volunteer bias due to the fact that females are over-represented in the UKB. Breast cancer is much more likely to occur in females than males, and sex-related participation bias plays a role in GWAS [6].

Table 3 provides additional comparisons of WGWAS and GWAS results for all ten phenotypes using WGWAS and GWAS effect sizes for all HapMap3 SNPs in the UKB. The first column shows the genetic correlation between the unweighted and weighted GWAS effect sizes, estimated through LD-score regression (see Methods). The correlation is positive and close to one for most phenotypes, but differs significantly from one for all, with the exception of BMI, height and age at first birth. The lowest congruence between weighted and unweighted SNP associations is found for Type 1 Diabetes (*rG* = 0.66) and Breast Cancer (*rG* = 0.81). We use the standard errors of WGWAS (GWAS) to estimate the effective sample size in columns 2 and 3 of the table (see Methods). Averaged over all phenotypes, the effective sample size shrinks from 337,534 in GWAS to 130,684 in WGWAS, a shrinkage of 61.2%. This implies that representative samples increase the power of GWAS, as the effective sample size shrinks when taking volunteer bias into account. Standard errors in WGWAS vis ’a vis GWAS are inflated by 32% to 87% on average (Table 3, column 4). Hence, when correcting genetic associations for selection bias using IP weighting, researchers face a bias-variance trade-off, as OLS is efficient in the absence of selection bias.

**Table 3:** Comparison of weighted and unweighted GWAS results. Comparisons use all UKB SNPs in HapMap3 (1,025,058 in total). The first column shows the genetic correlation between GWAS and WGWAS results, estimated through LD-score regression (see Methods). The second and third columns show the effective sample sizes (see Methods) of both methods. WGWAS increases standard errors by the percentage shown in column 4 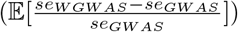. Column 5 shows the number of genomewide significant SNPs for each trait in GWAS, column 6 shows this in WGWAS. * values significantly different than one at Bonferroni-corrected level of 5% significance (*p <* 0.005)

| Phenotype | $r(\hat{\beta}_{GWAS}, \hat{\beta}_{WGWAS})$ | $N_{eff}^{GWAS}$ | $N_{eff}^{WGWAS}$ | Increase S.E.s | sig. hits<br>GWAS | sig. hits<br>WGWAS |
| --- | --- | --- | --- | --- | --- | --- |
| Age at First Birth | 0.976 (0.0128) | 139093 | 51949 | 71.3% | 30 | 3 |
| BMI | 0.992 (0.0052) | 372969 | 135238 | 76.7% | 1205 | 127 |
| Breast cancer | 0.813*(0.0341) | 376072 | 182605 | 32.2% | 51 | 8 |
| Drinks per Week | 0.936*(0.0188) | 265696 | 96008 | 83% | 23 | 4 |
| Self-rated health | 0.973*(0.0088) | 372714 | 136982 | 81.5% | 101 | 6 |
| Height | 0.993 (0.0032) | 374175 | 151328 | 60.6% | 5114 | 1453 |
| Physical activity | 0.866*(0.031) | 334570 | 123017 | 75% | 3 | 0 |
| Severe Obesity | 0.949*(0.0175) | 373834 | 136396 | 75.3% | 23 | 1 |
| Type 1 Diabetes | 0.66*(0.0566) | 373786 | 132605 | 87% | 69 | 37 |
| Years of Education | 0.988 (0.0062) | 392433 | 160707 | 63.8% | 331 | 49 |

The last two columns document how WGWAS decreases the number of genomewide significant SNPs relative to GWAS. Here, we only consider independent SNPs, identified through clumpling of WGWAS (GWAS) summary statistics (see Methods). For example, the number of genomewide significant SNPs in our BMI GWAS is 174, whereas it is 22 in the corresponding WGWAS. Such newly insignificant SNPs may indicate false positives in the current GWAS literature, but may also be a result of the increased standard errors that are a feature of WGWAS.

We use a Hausman test to calculate p-values for the null hypothesis that the effect sizes in weighted and unweighted GWAS are the same (see Methods). Here, we find that estimated SNP effects for breast cancer and type 1 diabetes are the most altered by weighting. For breast cancer and type 1 diabetes, we respectively find 365 and 40 independent loci with significantly altered SNP effects after weighting. 11 of these SNPs, tagging 3 independent loci, are not identified as genomewide significant for type 1 diabetes by GWAS, but are genomewide significant in WGWAS (Supplementary table 5). For example, rs341988 is insignificant for type 1 diabetes in GWAS (*β* = −0.0009 s.e. = 0.0007 *P* = 0.17), but is identified as genomewide significant in WGWAS (*β* = −0.004 s.e. = 0.0007 *P* = 1.26 · 10^*−*8^). The difference in these point estimates is significant (*P*_*H*_ = 4.05 · 10^*−*24^). Hence, for type 1 diabetes, selection bias takes the form of attenuation bias, and this results in missing several genomewide significant loci. A comparison of the manhattan plots for GWAS and WGWAS for type 1 diabetes clearly indicates that weighting alters which loci become significant and which ones become insignificant for this phenotype (Figure S4). Similarly, for breast cancer we find 3 SNPs, tagging one independent locus, that is insignificant in GWAS, but significant in WGWAS (Supplementary table 6). We followed up on these loci in the GWAS catalog (section S3). All four loci have not previously been identified as being associated with these phenotypes. Furthermore, two loci are entirely new to the literature: neither of these SNPs, nor any SNPs in linkage disequilibrium with these SNPs (*r*^2^ *>* 0.1, distance 500kb) have been reported in the GWAS catalog at *P <* 1 × 10^*−*5^ [22].

For the phenotypes drinks per week and physical activity we both find 1 SNP with a significantly altered effect size between WGWAS and GWAS, but these SNPs were not genomewide significant using any of the two methods. For the 6 other phenotypes, we find *no* evidence of any particular SNP having a genomewide significant difference in the effect size after volunteer bias correction. However, our qq-plots indicate that we are underpowered for these analyses, as they have a *λ* smaller than one (Figure S5). By contrast, the qq-plot of our GWAS with the IP weights as a phenotype shows an early liftoff (*λ* = 1.55), which suggests there are many SNPs throughout the genome whose observed frequencies may be affected by volunteer bias. In the remainder, we investigate how weighting of GWAS results affects various downstream statistics that rely on GWAS results.

### 2.2 SNP heritability estimates become larger after correcting for volunteer bias

We estimated SNP-based heritabilities, the proportion of genetic variation that can be explained by common SNPs, for all tested phenotypes using LD-score regression (see Methods) based on GWAS/WGWAS. In estimation, we use the effective sample sizes (summarized in Table 3) to account for the increased estimation error of WGWAS vis a’ vis GWAS [23]. The results are summarized in Table 4.

**Table 4:** SNP-based heritabilities for GWAS and WGWAS. SNP-based heritabilities were estimated using LD-score regression (see Methods). Heritabilities are shown here on the observed scale. The third column shows the p-value for the null hypothesis that the GWAS and WGWAS heritabilities are the same. The fourth and fifth columns show the intercept of the LD-score regression in GWAS and WGWAS, respectively. An intercept *>* 1 can be attributed to bias arising from population stratification [24].

| Phenotype | GWAS $h^2$ (SE) | WGWAS $h^2$ (SE) | P | GWAS Intercept (SE) | WGWAS Intercept (SE) |
| --- | --- | --- | --- | --- | --- |
| Age at First Birth | 0.1657 (0.0073) | 0.2135 (0.0143) | $1.28 \times 10^{-5}$ | 1.0347 (0.0096) | 1.0147 (0.008) |
| BMI | 0.2281 (0.0065) | 0.2381 (0.0091) | 0.14 | 1.127 (0.0152) | 1.033 (0.011) |
| Breast cancer | 0.0149 (0.0018) | 0.0267 (0.0029) | $1.11 \times 10^{-7}$ | 1.0183 (0.0079) | 0.9825 (0.007) |
| Drinks per Week | 0.0599 (0.003) | 0.0739 (0.0054) | $7.44 \times 10^{-4}$ | 1.0051 (0.0077) | 0.9852 (0.0064) |
| Height | 0.4235 (0.0189) | 0.4464 (0.0206) | 0.059 | 1.4785 (0.0345) | 1.1694 (0.0195) |
| Physical activity | 0.0281 (0.0019) | 0.031 (0.0044) | 0.408 | 0.9962 (0.0069) | 0.9933 (0.0069) |
| Self-rated health | 0.0972 (0.0029) | 0.125 (0.0052) | $9.35 \times 10^{-13}$ | 1.0522 (0.0103) | 1.0091 (0.0079) |
| Severe Obesity | 0.0416 (0.0022) | 0.0584 (0.0045) | $1.83 \times 10^{-6}$ | 1.0166 (0.0082) | 0.995 (0.0076) |
| Type 1 Diabetes | 0.0054 (0.0014) | 0.0432 (0.0035) | $1.63 \times 10^{-41}$ | 1.0194 (0.0074) | 0.9403 (0.0064) |
| Years of Education | 0.1482 (0.0052) | 0.1775 (0.0073) | $2.07 \times 10^{-9}$ | 1.1635 (0.0155) | 1.0531 (0.0113) |

For most phenotypes, correcting for volunteer bias by WGWAS results in substantial increases in the SNP-based heritability estimates. As for the SNP associations we explored in section 2.1, weighting matters the most for the heritability estimates of type 1 diabetes and breast cancer. For type 1 diabetes, the SNP-based heritability increases from 0.54% in GWAS to 4.32% in WGWAS, a large and highly statistically significant increase (*P* = 1.63 · 10^*−*41^). For breast cancer, the heritability almost doubles from 1.49% to 2.67% (*P* = 1.11 · 10^*−*7^). Most other phenotypes also see their heritabilities increased. For example, EA has a heritability of 14.8% in the UKB, but this increases to 17.8% when volunteer bias is taken into account (*P* = 2.07 · 10^*−*9^). Drinks per week, severe obesity, age at first birth and self-rated health also show substantial increases in estimated SNP heritabilities. This is consistent with phenotype-based selection (*scenario 1* in our simulations; Table 1). By contrast, Height, BMI, and Physical Activity do not show significant changes in heritability.

In LD-score regression, an intercept greater than 1 may be indicative of bias due to population stratification or cryptic relatedness [24]. For our *unweighted* GWASs, we find intercepts larger than 1 for years of education, BMI, height, self-rated health, and age at first birth as is common for these phenotypes [25–27]. As a consequence of weighting, the intercept moves closer to one, and becomes statistically indistinguishable from one for self-rated health and age at first birth. Hence, WGWAS may have the additional advantage of reducing bias arising from population stratification.

### 2.3 Volunteer bias affects gene tissue expression analysis

Gene tissue expression analysis is a popular tool for understanding the biological pathways through which genes may operate on a phenotype. We assessed the relevance of volunteer bias for such bio-annotations by conducting gene-set analyses using our WGWAS and GWAS summary statistics in MAGMA (implemented through the FUMA pipeline)[28, 29]. Here, we highlight the results for the breast cancer WGWAS and GWAS (Figure 1). For this phenotype, unweighted GWAS results show no evidence of genes expressed in any particular area of the body being significantly more associated with the likelihood of breast cancer. However, when estimating the same associations through WGWAS, we find that genes expressed in the fallopian tube, uterus, ovary, and breast mammary tissue are more likely to exhibit associations with breast cancer. Thus, correcting GWAS for volunteer bias may improve understanding of the pathways through which the genome influences a phenotype of interest.

**Figure 1:**
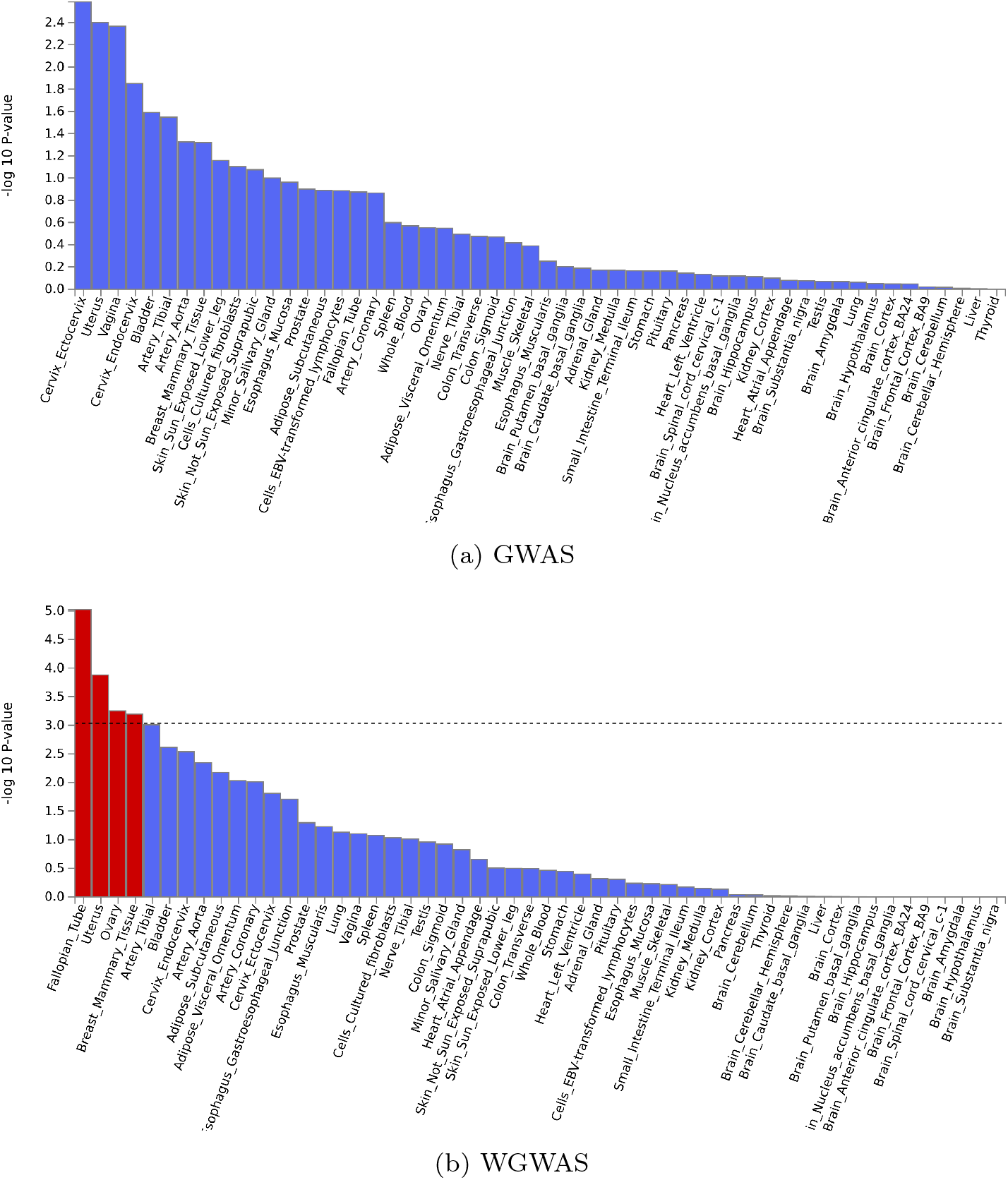
Gene-set analysis for Breast Cancer, estimated using MAGMA, for GWAS, and WGWAS that corrects for volunteer bias in the UKB

In supplementary material (Figure S7 to Figure S14), we show MAGMA gene tissue expression analyses for the 9 other phenotypes. We find several phenotypes for which significant areas of expression are found in GWAS, but not in WGWAS, namely age at first birth, BMI, self-reported health, and physical activity, suggesting that such findings might possibly be spurious and driven by volunteer bias.

## 3 Discussion

Our analyses highlight the drawbacks of non-random, volunteer-based sampling for GWAS and subsequent downstream genetic analyses. Contrasting WGWAS with GWAS results for ten phenotypes, we demonstrated that in GWAS volunteer bias results in (i) missing genomewide significant loci for type 1 diabetes and breast cancer, (ii) attenuated effect sizes and missing heritability for various health-related and behavioral phenotypes, and (iii) biased gene-tissue expression findings. Our results suggest that the need to correct GWAS results for volunteer bias through IP-weighting (i.e. WGWAS) is phenotype-specific. Phenotypes where weighting altered results substantially are disease-related (e.g. Type 1 diabetes, breast cancer), related to socioeconmic status (e.g. educational attainment) or related to health behavior (e.g. drinks per week). By contrast, height is an example of a phenotype where weighting made a relatively minor difference. Although weighting still altered results for height, practitioners may wish to opt for GWAS, rather than WGWAS for such a phenotype, because of the bias-variance tradeoff, which increases the standard errors of WGWAS vis ’a vis GWAS.

The focus here was on the UKB for which we estimated IP weights [3]. Many other GWAS cohorts are volunteer-based and will suffer from similar, but distinct, forms of volunteer bias. Our results suggest that such volunteer biases need to be taken seriously, and should be corrected for. GWAS consortia may wish to ensure that weights are available for all volunteer-based cohorts included in their GWAS. Such IP weights can be estimated by comparing the genotyped data set to a source of representative data (e.g., Census data or administrative data), provided that both data sets have a sufficient number of (close to) identically measured variables in common. Further, in the design of a new data set, it is essential that as many variables as possible are collected that are shared with a source of representative data to ensure that IP weights can be precisely estimated. Our results suggest that IP weighting is sufficient to capture a substantial degree of volunteer bias in genetic association results. WGWAS increases standard errors, but is also likely to increase effect sizes, such that power need not be reduced. Further, WGWAS reduces the effective sample size of a cohort, which should be taken into account when meta-analyzing multiple cohorts.

Our results provide insights into the effects of volunteer bias on GWAS, but drawbacks remain. The IP weights we use to correct for volunteer bias may suffer from omitted variable bias, since the model that was used to create them only includes variables that the UKB and UK Census safeguarded microdata have in common. These variables mostly capture socioeconomic status, demographics and self-reported health. However, it is possible that other variables that relate to UKB volunteering are missing, e.g., personality characteristics. Nonetheless, these weights have been shown to capture an average of 78% of volunteer bias in phenotype-phenotype associations [3], and we therefore expect that the genomic results presented here can indeed be interpreted as being representative of the population from which the UKB was sampled.

## Data Availability

UKB data can be accessed upon request for research projects that have obtained the necessary approval. These requests can be submitted through https://www.ukbiobank.ac.uk/

## Acknowledgements

Research reported in this publication was supported by the National Institute On Aging of the National Institutes of Health (RF1055654 and R56AG058726), the Dutch National Science Foundation (016.VIDI.185.044), and the Jacobs Foundation. This research has been conducted using the UK Biobank Resource under Application Number 55154. We thank participants at the 2021 and 2022 BGA annual meeting, 2021 and 2022 ASHG conference, the 2021 Integrating Genetics and Social Science Conference, and the 2022 European Social Science Genetics Network Conference for their feedback and comments. We also thank Michel Nivard, Ronald de Vlaming and Hyeokmoon Kweon for their kind feedback and valuable comments.

## 4 Methods

### 4.1 Data

#### 4.1.1 UK Biobank (UKB)

The UKB is a cohort of 503,317 individuals collected between 2006 and 2010 at 22 assessment centres spread out across Great Britain. Potential participants were identified through the registry of the National Health Service, which covers virtually the whole UK population. Individuals living in proximity to an assessment centre and aged 40 to 69 at the start of the assessment period (which varies per assessment centre) received an invitation to participate by post. This UKB-eligible population consists of 9,238,453 individuals who received an invite, such that the overall acceptance rate was 5.45%.

Figure S18 summarizes our sample selection criteria for the UKB. We drop individuals that were not included in the genetic subsample, and restrict the UKB to individuals who identified as “white British” and were of genetic European ancestry, as most published work with the UKB genetic data makes this sample restriction [e.g. 25, 26, 30]. We also drop respondents that did not meet the standard requirements regarding genetic data quality control (see next subsection). Last, we dropped 6,292 respondents (1.6%) for whom IP weights could not be estimated, typically because of missing variables (see ref [3] for detail)

#### 4.1.2 Genetic data in the UKB

Genetic data collection on UKB participants has been extensively described elsewhere [31]. We restrict our sample to those of white British ancestry, as defined by a PC analysis conducted by Bycroft et al.[31] As is the standard in GWAS analyses, we only keep UKB participants that were sufficiently densely genotyped: we drop individuals that have missing values at more than 2% of all SNPs measured in UKB (6,118 participants in total). We also drop those with outlying heterozygosity values (mean +*/*− 3 std. deviations of the heterozygosity distribution observed in the data; 2,279 participants in total). Furthermore, we drop individuals for whom their reported sex does not match with their sex as inferred from their measured genome (296 in total), as such mismatch may point towards sample contamination or sample mix up. We focus on a genotyped sample that is approximately independent by keeping only one individual from each group of first-degree relatives. The individual that is kept is the one with the least missingness in their genetic data. As a result, we drop 18,736 respondents from the sample.

We conduct our analyses on autosomal SNPs which are in HWE (*p >* 1 × 10^*−*6^), with *MAF >* 0.01, and which are missing in less than 2% of all included respondents. For reasons of computational feasibility, we only select 1,025,058 of these SNPs that were also available in the HapMap3 reference panel.

### 4.2 GWAS on the IP Weights

We estimate a GWAS using the IP weights as a phenotype by fitting a linear model in PLINK, restricting to our quality-controlled set of HapMap3 SNPs. Independent hits were assessed through PLINK’s clumping algorithm (*R*^2^ ≥ 0.1, LD-window of 250kb). SNP-based heritability was estimated using LD-score regression [24].

### 4.3 Regular GWAS and WGWAS

For each phenotype, we estimate GWAS associations for all HapMap 3 SNPs that were available in the UKB data. We fit the following model:

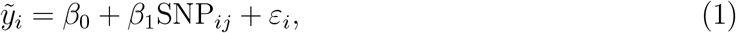

where 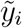 is the estimated residual of the phenotype from an auxiliary regression which fits *y*_*i*_ on a set of variables that may confound the relationship between *SNP*_*j*_ and *y*. These variables are genetic sex, the first 20 principal components, gene batch fixed effects, and a dummy for individual i’s birth year cohort (5-year bins) capturing the effects of ageing on *y*_*i*_. SNP_*ij*_ is individual *i*^*′*^*s* allele count at the *i*th SNP.

We estimate two GWASs for each phenotype: (1) a regular GWAS, which estimates SNP associations using the above model by OLS, and (2) an inverse probability weighted GWAS (WGWAS), which estimates the above model using the IP weights as estimated in [3] using a subsample of 2011 UK Census microdata as data representative of the UKB’s sampling population. For WGWAS, 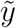 was residualized using the same weights in the auxiliary regression. We estimate heteroskedasticity-robust (White) standard errors for both GWAS and WGWAS. Both GWAS and WGWAS were estimated in R.

Approximately independent SNPs were assessed through PLINK’s clumping algorithm (*R*^2^ ≥ 0.1, LD-window of 250kb).

### 4.4 Comparing GWAS and WGWAS results for known top hits

Known top hits were selected from publicly available GWAS results that did *not* include the UKB as part of their discovery sample (See supplementary table 2). To obtain top hits that were approximately independent, we clumped these results (*R*^2^ ≥ 0.1, LD-window of 250kb). Top hits were selected by only selecting SNPs with cutoff *p <* 10^*−*5^.

### 4.5 Testing for significant differences in WGWAS and GWAS associations

P-values (denoted *P*_*H*_) that test the null hypothesis 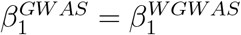 are estimated using the Hausman test statistic: 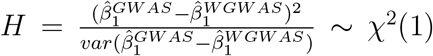. In this expression we use 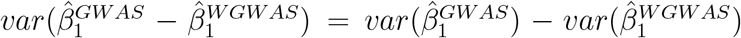, given that 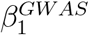 is estimated efficiently under the null [32, 33].

### 4.6 Determining the effective sample sizes of GWAS and WGWAS

The effective sample size aids to understanding how much non-representativeness dilutes the power of GWAS results, and are a crucial input into the LD-score regressions (see next section). We calculate the effective sample size for each SNP, given by

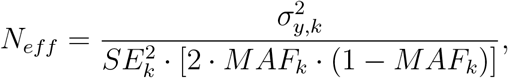

with *k* referring to either the unweighted or IP weighted sample statistic [23]. 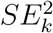 is given by the standard error of the SNP as determined by unweighted or IP weighted GWAS, respectively. For each phenotype, the effective sample size as averaged over all SNPs is reported.

### 4.7 SNP-based heritabilities and genetic correlations

We use LD-score regression to estimate the genetic correlation and SNP-based heritabilities for GWAS and WGWAS [24, 34]. GWAS and WGWAS summary statistics were prepared using the munge sumstats.py function. Our estimates of *N*_*eff*_ were used as the parameter for the sample size when preparing the summary statistics for both GWAS and WGWAS.

To evaluate whether our SNP-based heritabilities differed for GWAS and WGWAS, we construct the following Z-statistic:

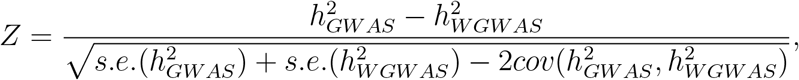

where we compute the covariance through

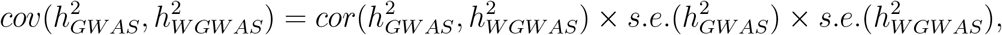

estimating 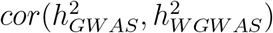 as the value of the intercept from the cross-trait LD-score regression on the weighted and unweighted GWAS results [23, 34].

## Footnotes

1 When two variables *X* and *Y* independently cause a third variable *Z*, collider bias leads to biased estimates of the association between *X* and *Y* conditional on *Z* [4, 11]. In this case, variables associated with volunteering (e.g., education or health) act as colliders, such that a spurious association between these variables may arise in the volunteer-based data.

2 The issue of non-random selection of the intended study population into the final sample is often termed selection bias [4, 5, 11, 12], participation bias [6, 7], volunteer bias [11, 13], or ascertainment bias [14], and it arises due to non-random selection from the population of interest *into* the actual sample. Perhaps confusingly, the term “ascertainment bias” is sometimes also used to describe situations where a dataset is representative of its intended study population, but this intended study population is not the same as the population of interest [15, 16].

3 Other large genotyped samples of similar (or larger) sample sizes, e.g., those of 23andMe, suffer from comparable volunteer biases due to self-selection of respondents.

4 An earlier paper estimated IP weights using the Health Survey for England [20], which has a smaller sample size (N = 7,721), fewer variables (6), and no geographic identifiers.

5 For example, UKB respondents have received an average 13.8 years of education (SD=4.91), which reduces to 13.0 years after weighting (SD=5.00).

