## Supplementary Notes and Figures for "Correcting for volunteer bias in GWAS uncovers novel genetic variants and increases heritability estimates"

### Contents

**S1 Coding of phenotypes** **2**

**S2 GWAS on the inverse probability weights** **4**

**S3 Follow-up of new loci found by WGWAS in the GWAS catalog** **6**

**S4 Supplementary figures** **8**

### S1 Coding of phenotypes

#### S1.1 Age at first birth

Age at first birth was assessed for females only and was derived from data field 2754 ("How old were you when you had your FIRST child?"). Respondents could indicate a numerical value, or could answer "Do not remember" or "Prefer not to answer", in which cases the variable was coded as missing.

#### S1.2 BMI

We used measured BMI as reported in data field 210001.

#### S1.3 Breast Cancer

Diabetes was derived from data field 40006 (Type of cancer: ICD10). Cases of breast cancer were defined by codes C50.0-C50.9.

#### S1.4 Diabetes - Type 1

Diabetes was derived from data field 41202 (Diagnoses - main ICD10), and, 41204 (Diagnoses - secondary ICD10). Cases of type 1 diabetes were defined by codes E10.0-E10.9.

#### S1.5 Drinks per week

Drinks per week was constructed from data field 1568 (average weekly red wine intake), 1578 (average weekly champagne plus white wine intake), 1588 (average weekly beer plus cider intake), 1598 (average weekly spirits intake), 1608 (average weekly fortified wine intake), and 5364 (average weekly intake of other alcoholic drinks). These values were self-reported. On each question, respondents could indicate "Do not know" or "Prefer not to answer". We coded values for respondents who filled out these options on any of these questions as missing, with the exception of data field 5346, for which we put a value of zero. Drinks per week was then defined as the sum of all these data fields as reported during the first non-missing wave.

#### S1.6 Height

We use measured standing height (in cm) as reported in data field 50.

#### S1.7 Health (self-reported)

For self-reported health, we use data field 2178 ("In general how would you rate your overall health?"). Respondents could answer on a likert scale of 1-4 (1: Excellent, 2: Good, 3: Fair, 4: Poor). We inverted this likert scale such that a higher value implies better self-reported health. Respondents could also indicate "Do not know" or "Prefer not to answer". These instances were coded as missing.

#### S1.8 Physical Activity

We measure physical activity as the sum of duration of moderate physical activity (data field 894) and vigorous physical activity (data field 914). Both frequencies were self-reported and measured as *minutes per day*, we converted this to minutes per week by multiplying by 7. Next, we converted this measure to the metabolic equivalent of moderate and vigorous activity combined, by multiplying moderate activity by 4, vigorous activity by 8, and taking the sum [36].

#### S1.9 Severe Obesity

Severe obesity was derived from data field 41202 (Diagnoses - main ICD10), and, 41204 (Diagnoses - secondary ICD10). Cases of severe obesity were defined by codes E66.0-E66.9.

##### S1.10 Years of Education

For years of education, we follow the coding procedure as in the most recent GWAS for educational attainment [37].

#### S2 GWAS on the inverse probability weights

In supplementary table 4, we list the 7 genomewide significant SNPs foundin our GWAS on the IP weights, as well as suggestive top hits ($P \leq 5 \cdot 10^{-5}$, 409 approximately independent SNPs in total). Researchers who study these loci in the UKB, or who find that these loci pop up in hypothesis-free approaches (e.g. GWAS) are advised to use an IP weighting procedure to investigate whether their results are not driven by volunteer bias.

Analyzing all GWAS results of our GWAS on the IP weights, the quantile-quantile plot of the p-values shows an early lift-off (Figure S2, $\lambda = 1.55$), which implies that volunteer bias may potentially impact associations of genetic markers across the whole genome. We assessed the genetic overlap between voluntary participation behavior and various phenotypes by estimating the genetic correlation between our GWAS on the IP weights and various phenotypes of interest. To this end, we estimated genetic correlations between the summary statistics of our IP weight GWAS and various publicly available GWAS results. The sources of these publicly available GWAS results are summarized in supplementary table 3. Figure S3 shows these genetic correlations: Our GWAS on the IP weights correlates mostly with Educational attainment (rG = -0.7114 (0.025)) and Age at First Birth (rG = -0.6978 (0.0293)). Since our IP weights are *inversely proportional* to UKB participation behavior, this implies that those with a genetic propensity towards education and giving birth at a later age are more likely to volunteer for UKB participation. Further, our GWAS on the IP weights shows substantial overlap with genes that associate with various measures of participation into optional modules of the UKB. These genetic correlations are again negative, which implies that those with higher IP weights are indeed less likely to volunteer for scientific data sets. Using these genetic correlations, we also find that volunteering into the UKB is positively associated with subjective well-being and height, and negatively associated with BMI and other weight-related phenotypes, Depression, and smoking behavior. These patterns are consistent with healthy volunteer bias.

We investigated the 7 top hits for UKB participation in further detail. Moving beyond HapMap3 SNPs only, we re-estimated the GWAS on the IP weights for all SNPs found in the UKB that were in linkage disequilibirum ($R^2 > 0.1$ and within a 500 kb window size) with these top hits. Figure S15 maps these areas of the genome. We obtained data on SNP-trait associations from the GWAS catalog, which has collected over 400,000 SNP-trait associations at the moment of writing [22]. We only include genomewide significant findings from the catalog ($P < 5 \cdot 10^{-8}$). In Figure S15 SNPs, that have been found to associate with any other trait as reported in the catalog are annotated as such. For example, Figure S15a shows that the lead SNP rs4399146 on chromosome 1 significantly associates with our IP weights, and that SNPs in strong LD with this SNP have been reported to associate with high density lipoprotein cholesterol, total blood protein, platelet count, and red blood cell distribution width. For the other 6 SNPs, we find that they tag loci that have been reported to relate to educational attainment, alcohol consumption, hypothyroidism, leukocyte count, lymphocyte count, autoimmune disease, and intelligence.

To assess whether these 7 top hits were located in loci with more or less pleiotropy than average, we used global pleiotropy estimates based on 4,155 GWASs estimated by Watanabe et al [39]. Their study subdivides the genome in 3,362 independent loci groups, and measures loci-specific pleiotropy by the number of trait domains (i.e. grouped phenotypes) for which prior GWAS studies have found genomewide significant SNPs within these loci. To assess the pleiotropy of our 7 loci significantly associated with the IP weights, we merged our clumped SNPs to the number of associated trait domains within their loci. The mean number of associated trait domains in non-significant results (here defined as $P > 5 \times 10^{-5}$) was 2.57, whereas the mean was 3.25 for genomewide significant hits. A t-test to compare these means reveals that the difference is not significant (P=0.671). It is possible that some of these loci suffer from phenotype-based selection, which results in attenuation bias and hence a lower likelihood of pleiotropy, whereas other loci suffer from phenotype-genotype-based selection, which results in a higher likelihood of false positives and hence a higher likelihood of pleiotropy.

### S3 Follow-up of new loci found by WGWAS in the GWAS catalog

Using WGWAS, we found 3 independent loci that are genomewide significant for type 1 diabetes, with associations that differed significantly ($P_H < 5 \cdot 10^{-8}$) from their GWAS counterparts. For breast cancer, we found 1 such new independent locus. To assess whether these loci were tagged in other GWASs, we proceeded as follows. We obtained data on SNP-trait associations from the GWAS catalog, which has collected over 400,000 SNP-trait associations at the moment of this writing.[22] We only include genomewide significant findings from the catalog ($P < 5 \cdot 10^{-8}$).

To assess whether the novel loci we uncovered in WGWAS were reported as genomewide significant elsewhere, we considered our lead SNP (given by the lowest p-value in the region) and re-estimated the WGWAS on the trait for *all* SNPs that were in linkage disequilibrium with this lead SNP, and were available in the UKB (not just HapMap3 SNPs). Figure S16 and Figure S17 show zoomed in Manhattan plots around these lead SNPs for type 1 diabetes and breast cancer, respectively. We annotated each SNP with the traits for which significant associations were reported in the GWAS catalog, if any. As can be seen, none of the new SNPs we found tag loci that were previously reported for type 1 diabetes or breast cancer respectively. Thus the SNPs we identified are novel. rs17186868, found to associate with type 1 diabetes in WGWAS, is in strong linkage disequilibrium $R^2 > 0.9$ with a lead SNP that is associated with BMI-adjusted waist circumference, and in weaker linkage disequilibrium with lead SNPs associated with body height and BMI-adjusted hip circumference. rs12522568, found to associate with type 1 diabetes in WGWAS, shows some evidence of being in linkage disequilibrium with a lead SNP for adolescent idiopathic scoliosis.

#### S4 Supplementary figures

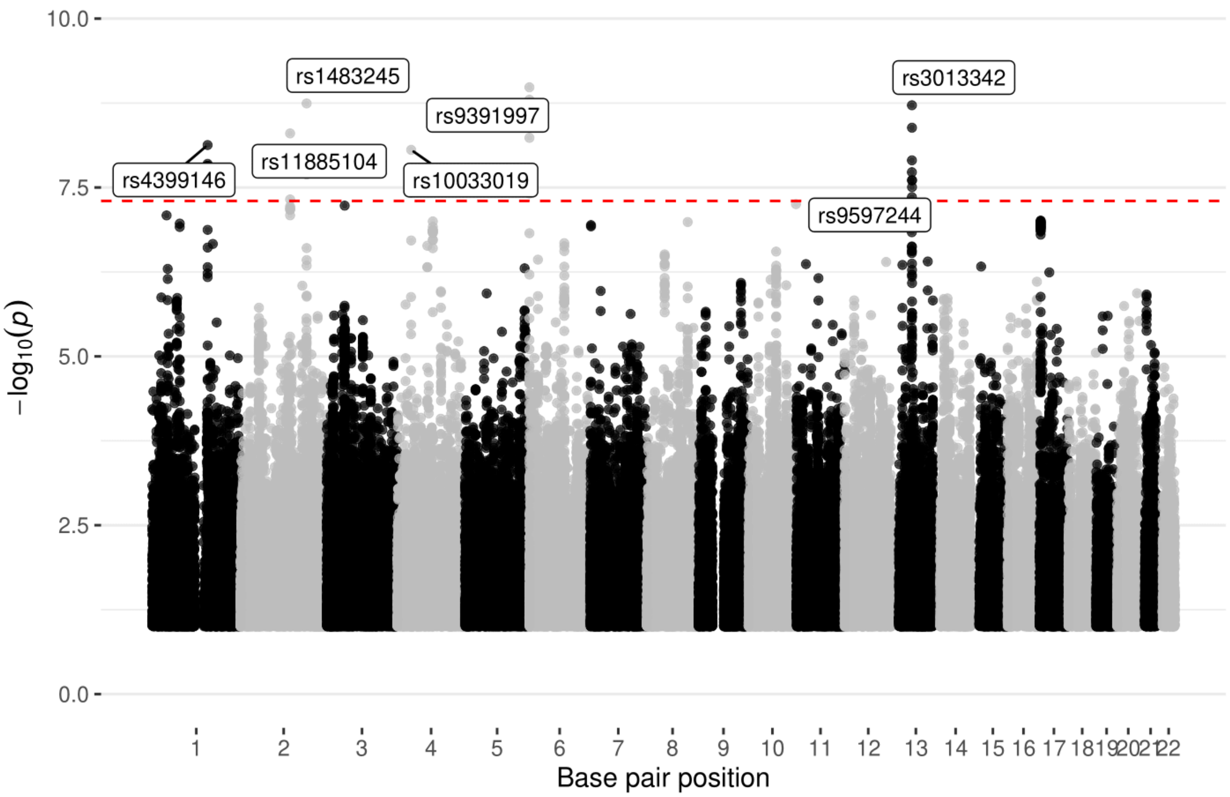


Figure S1: **Manhattan plot for the GWAS on the inverse probability weights.** The p-values are displayed on the y-axis on a $-log_{10}$ scale. The red line marks the genomewide significant threshold ($P = 5 \times 10^{-8}$). Approximately independent genomewide significant SNPs were assessed through clumping ($R^2 = 0.1$, window size 250kb). These top hits are annotated.

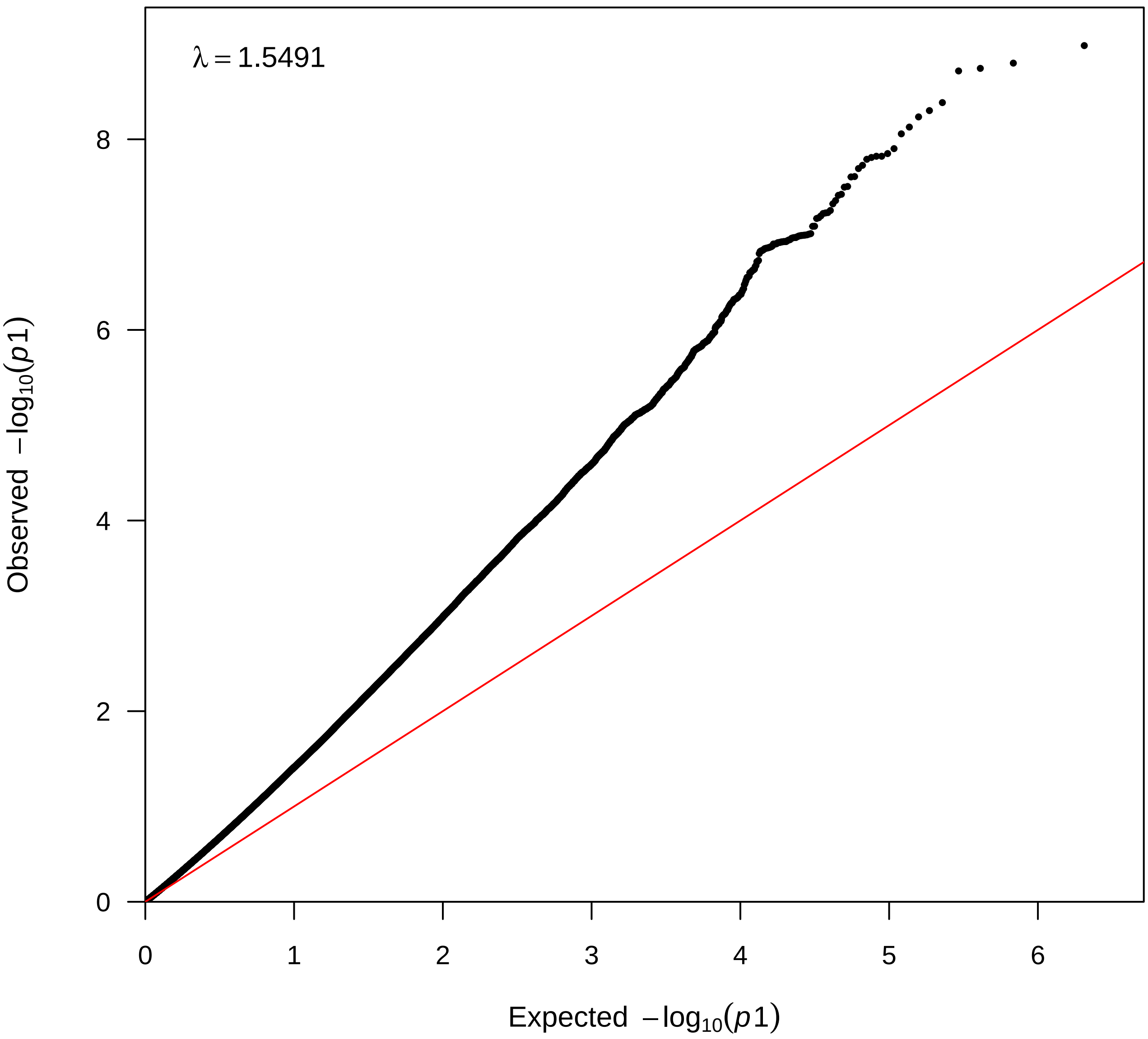


Figure S2: Quantile-quantile plot for the GWAS on the inverse probability weights. $\lambda$ refers to the genomic inflation factor [35].

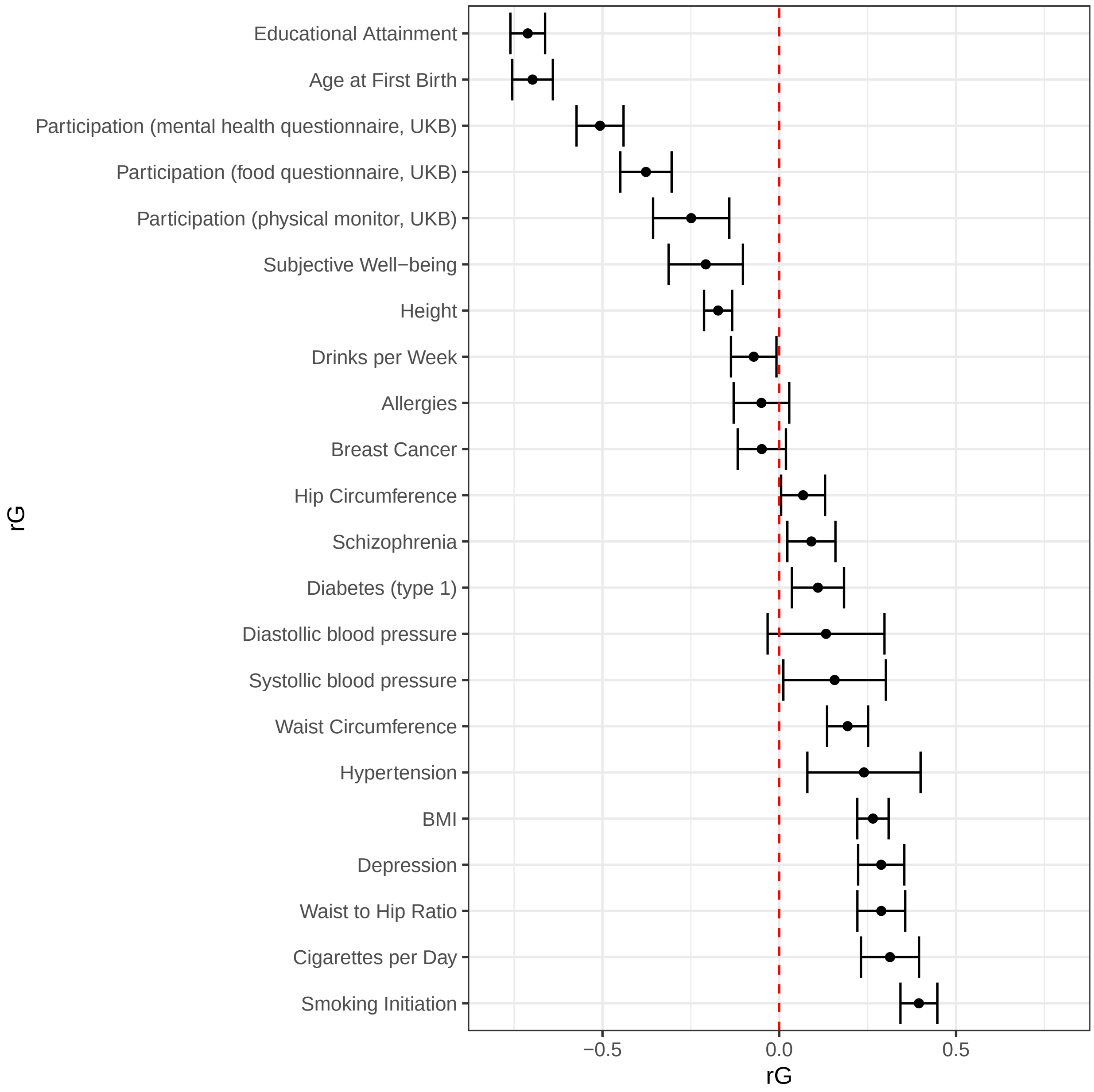


Figure S3: Genetic correlations between GWAS results on the inverse probability weights and various phenotypes

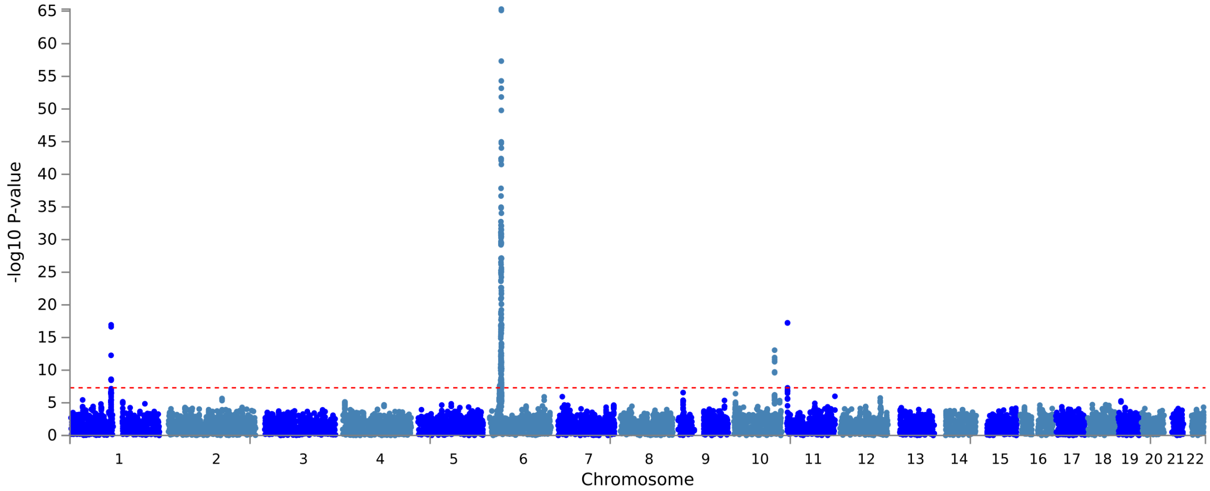


(a) GWAS

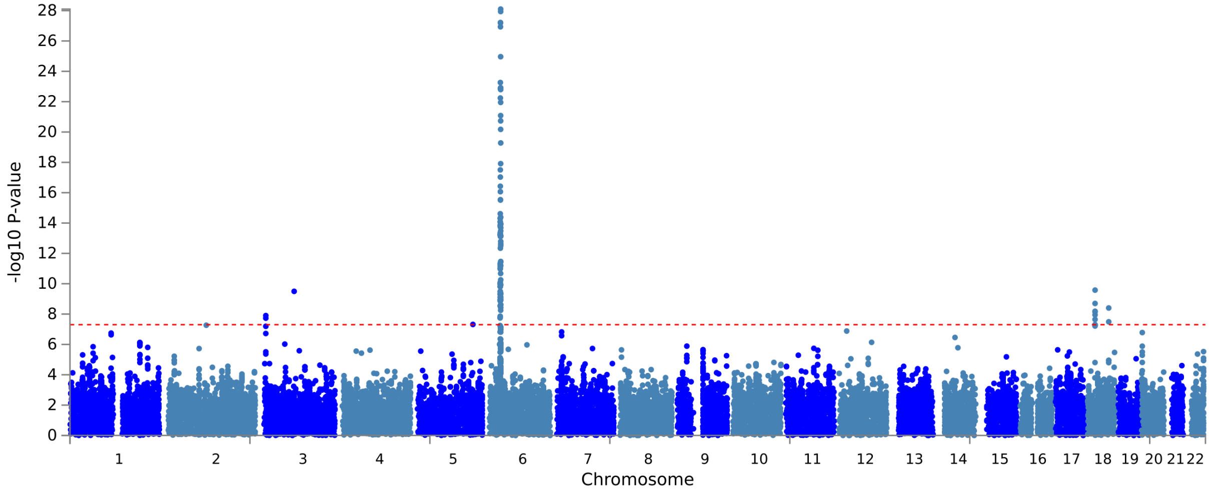


(b) WGWAS

Figure S4: Manhattan plot of GWAS and WGWAS results for type 1 diabetes

Figure S5: QQ plots of p-values which test for the difference between SNP associations estimated by GWAS and WGWAS for various phenotypes. $\lambda$ refers to the genomic inflation factor [35].

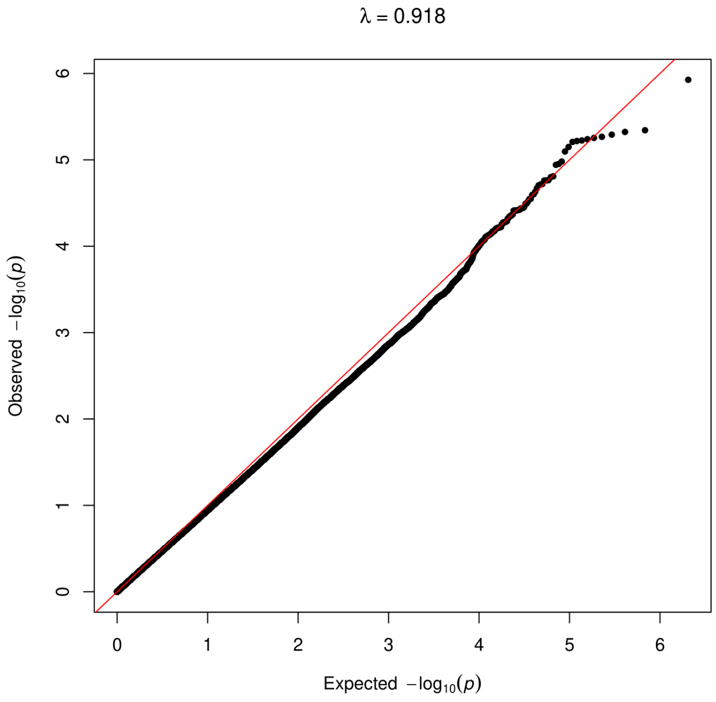


(a) AgeFirstBirth

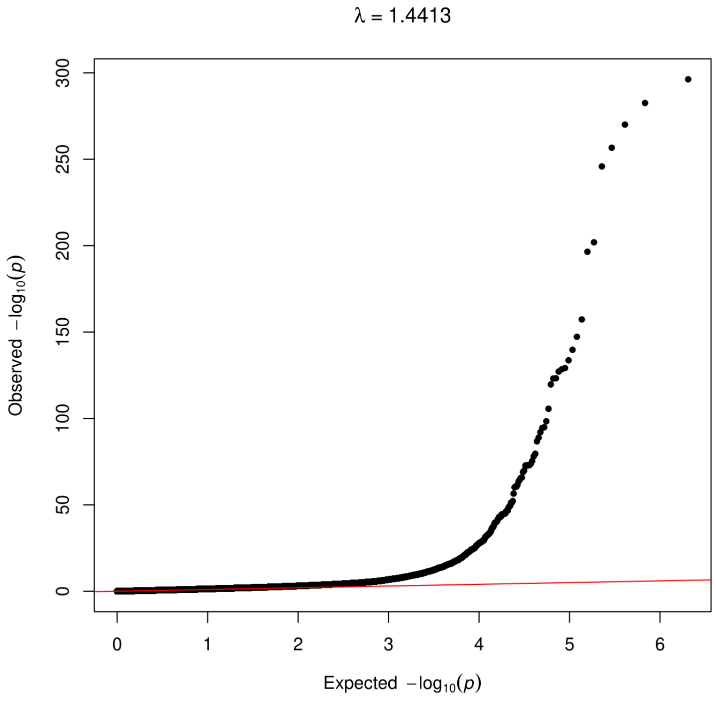


(b) Breast Cancer

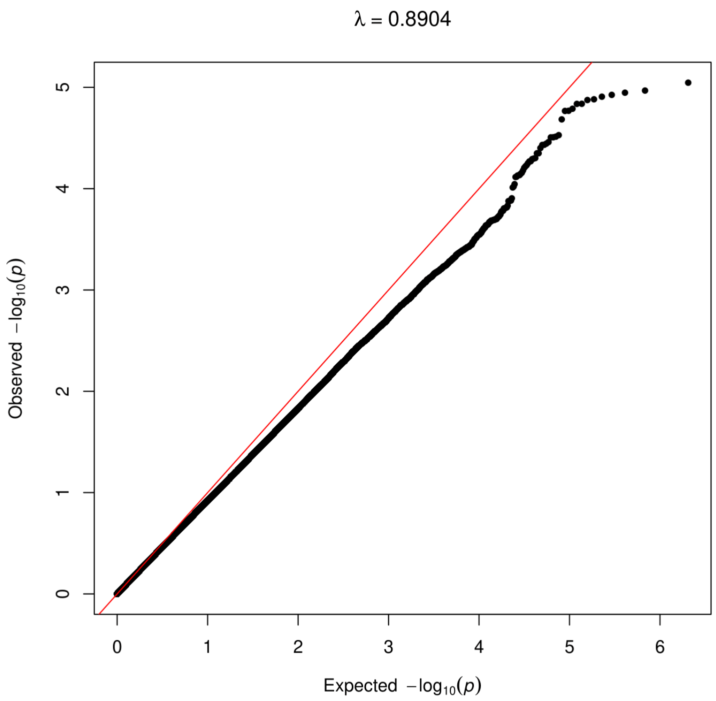


(c) BMI

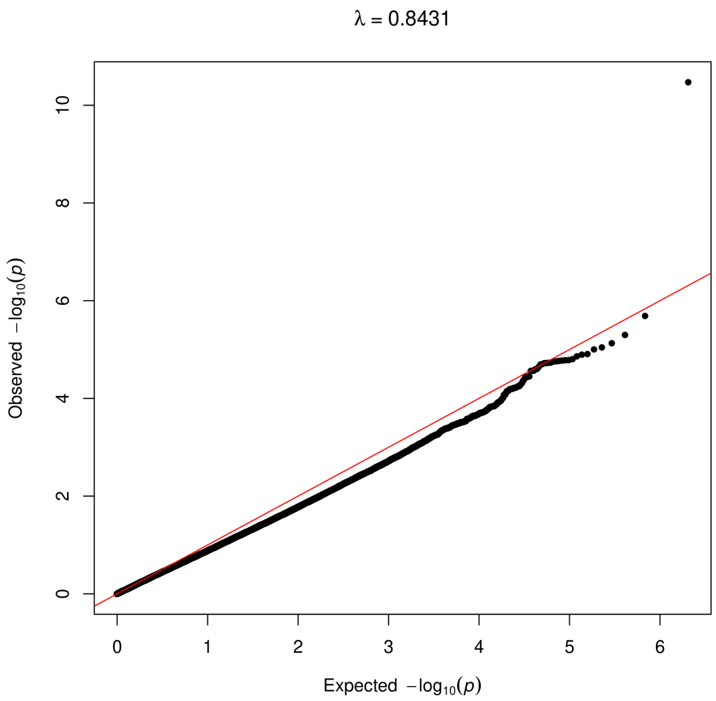


(d) Drinks Per Week

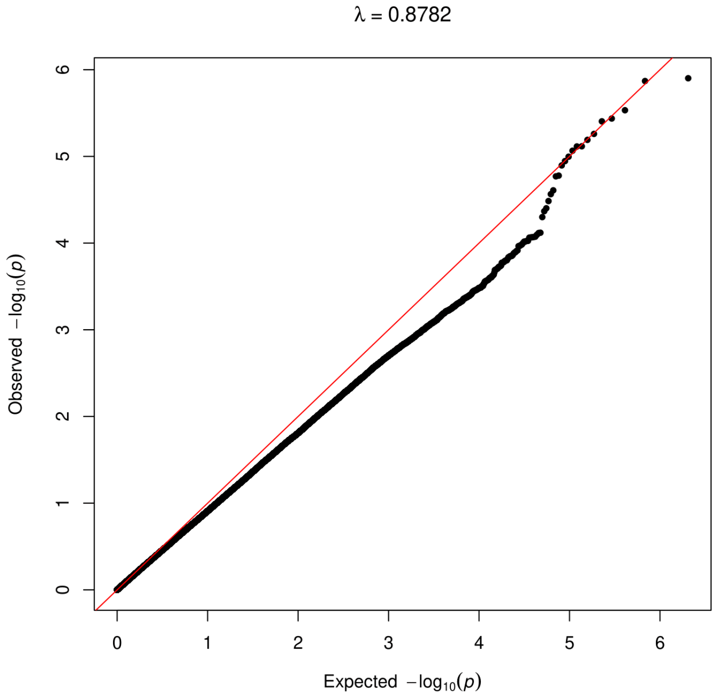


(e) Self-rated health

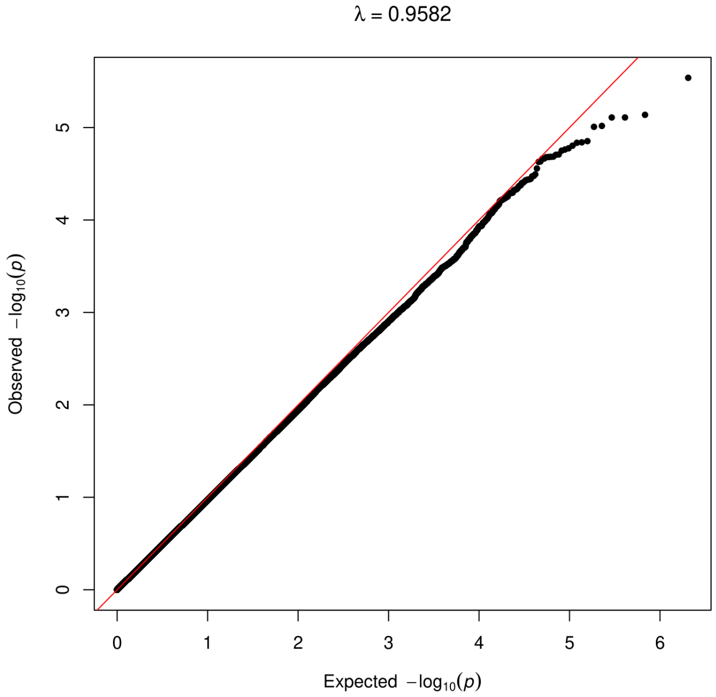


(f) Height

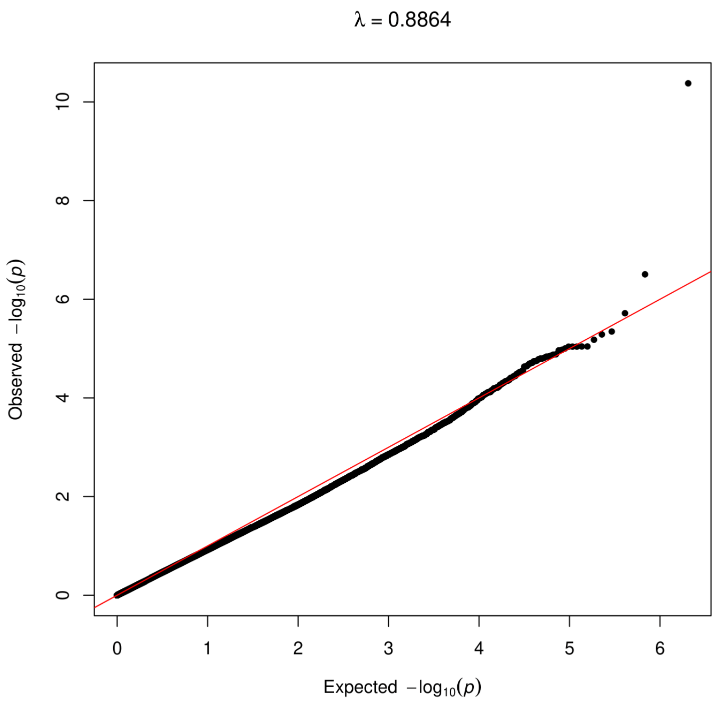


(g) Physical Activity

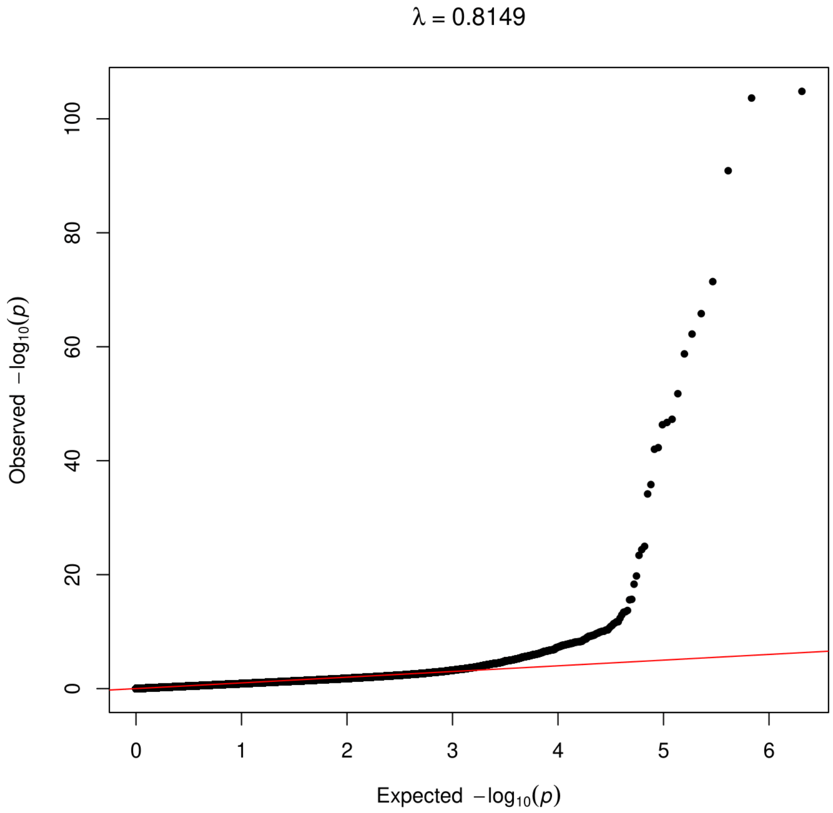


(h) Type 1 Diabetes

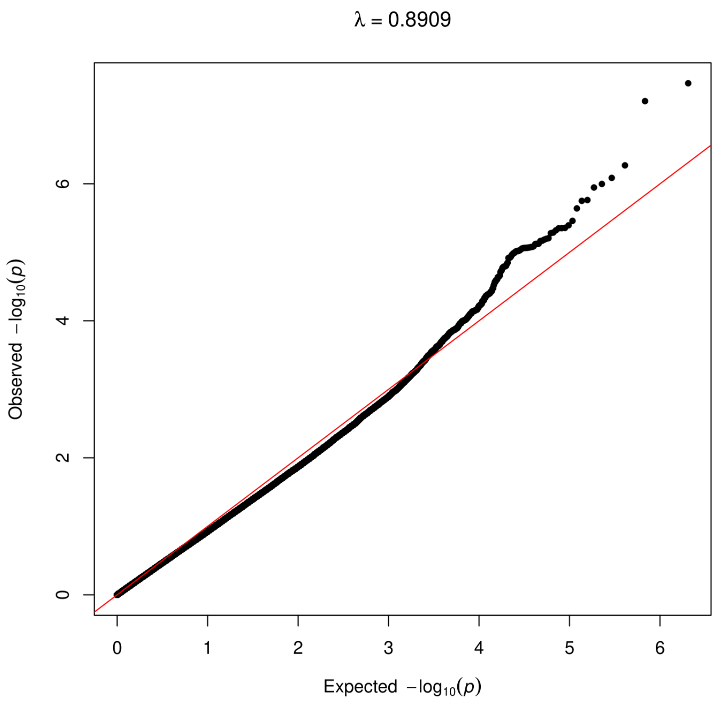


(i) Severe Obesity

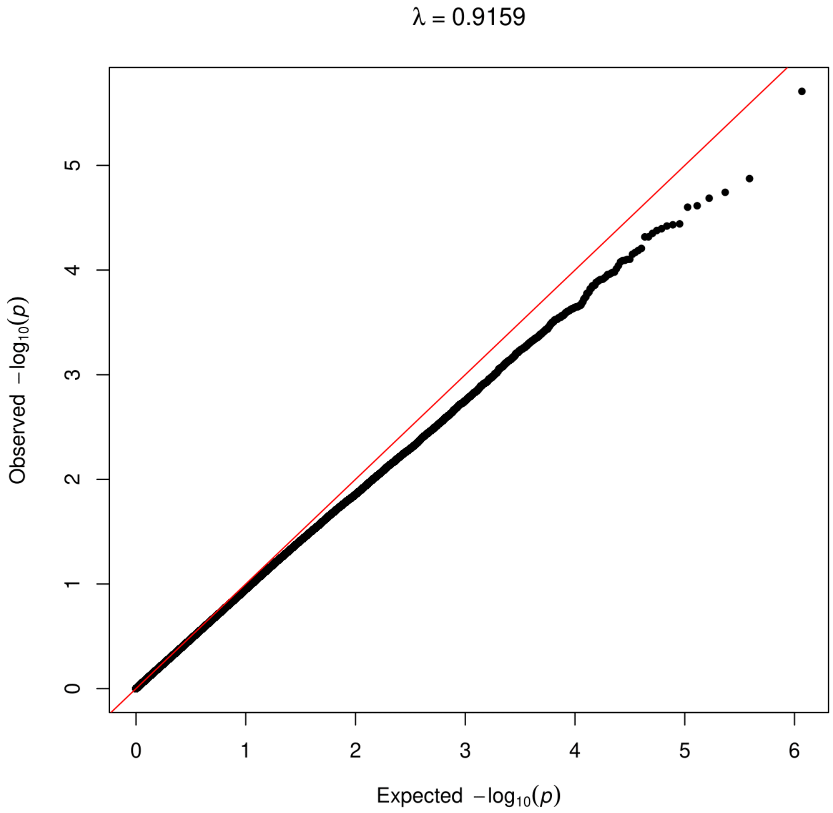


(j) Years of Education

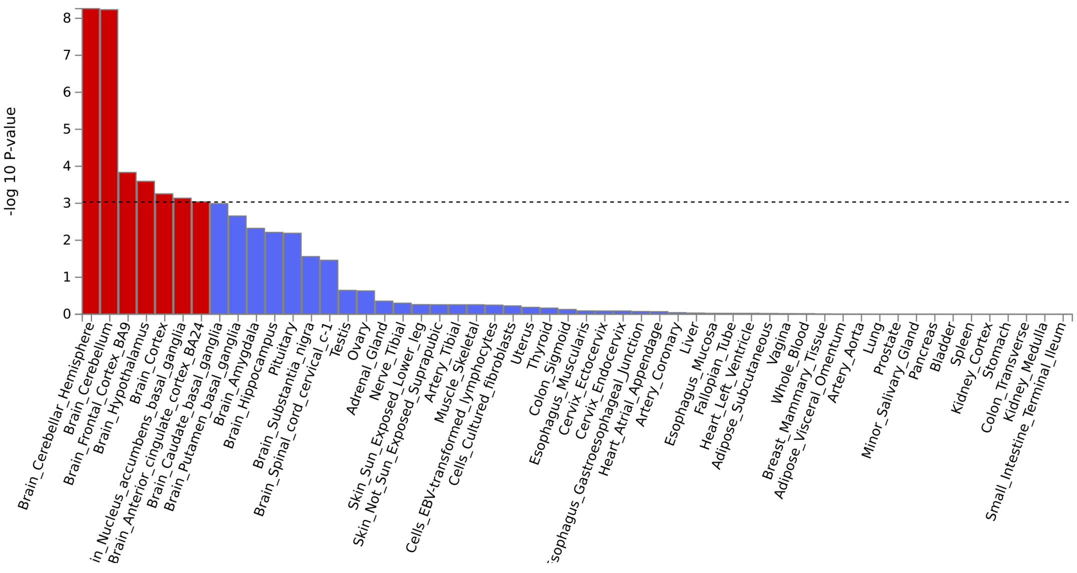


(a) AFB GWAS

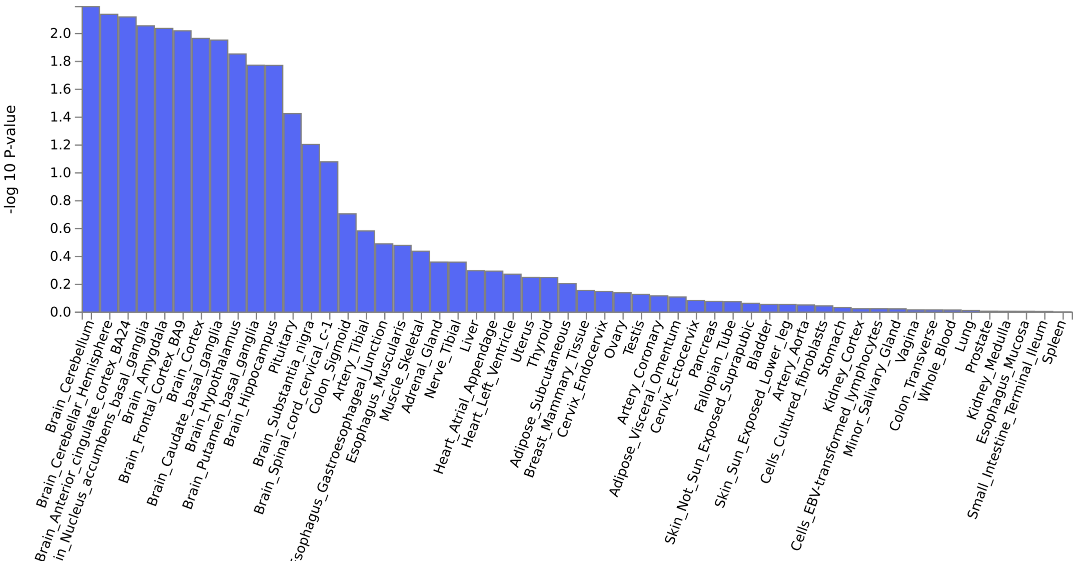


(b) AFB WGWAS

Figure S6: Gene tissue expression analysis estimated through MAGMA (implemented in FUMA) using GWAS/WGWAS results for Age at first birth

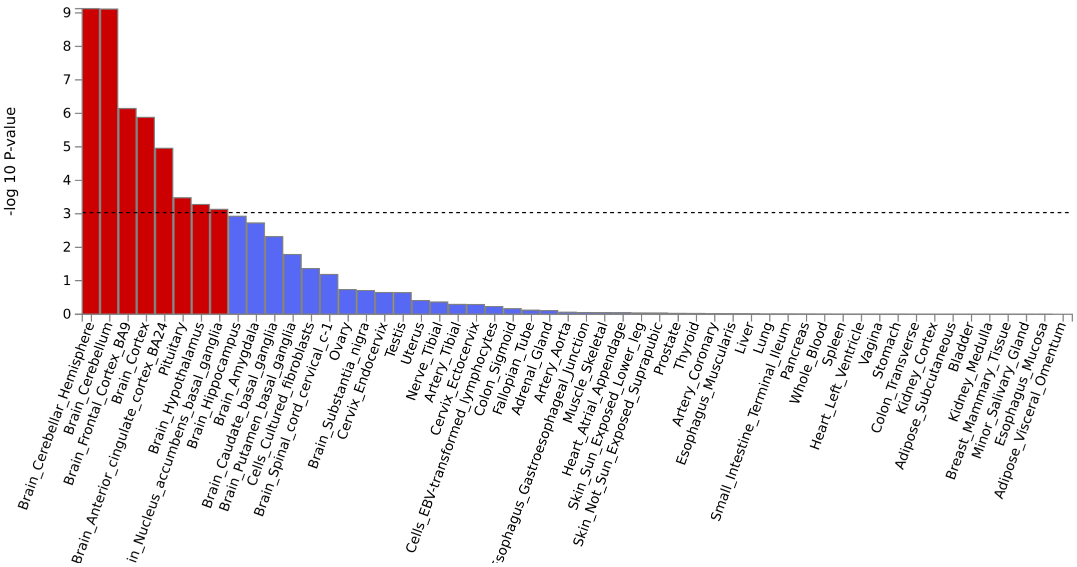


(a) BMI GWAS

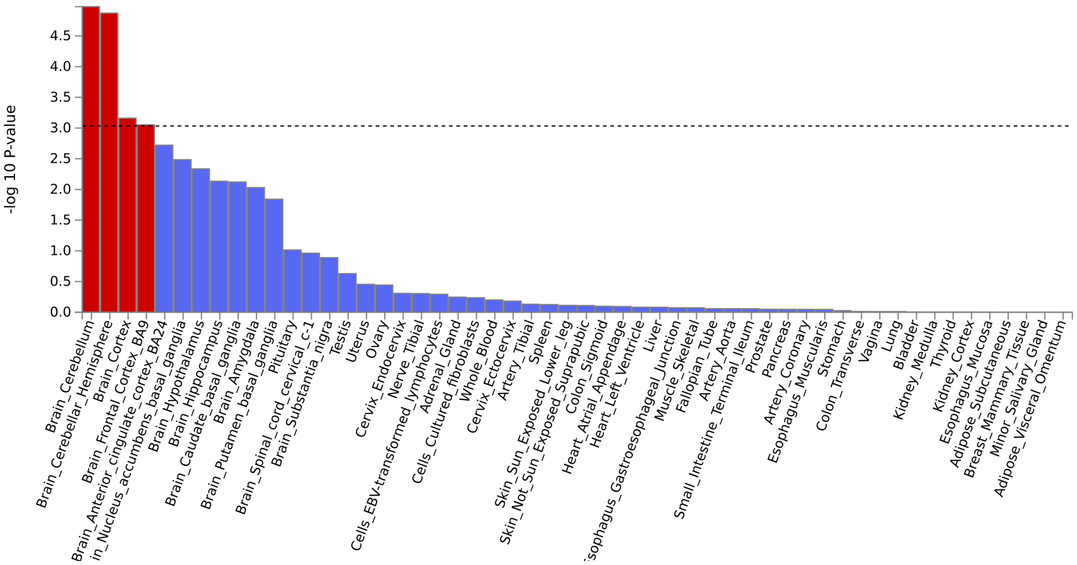


(b) BMI WGWAS

Figure S7: Gene tissue expression analysis estimated through MAGMA (implemented in FUMA) using GWAS/WGWAS results for BMI

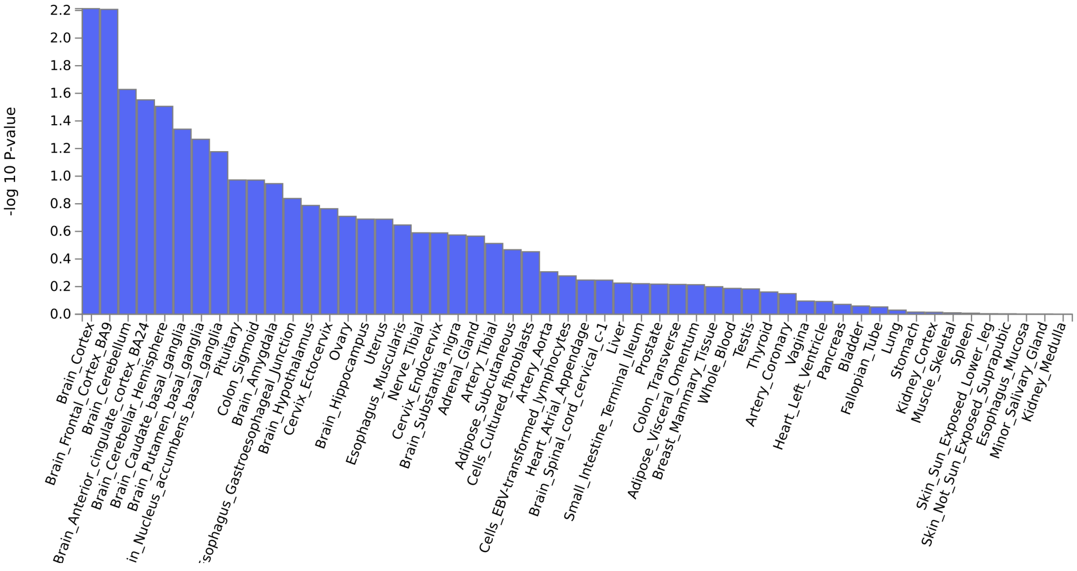


(a) Drinks Per Week GWAS

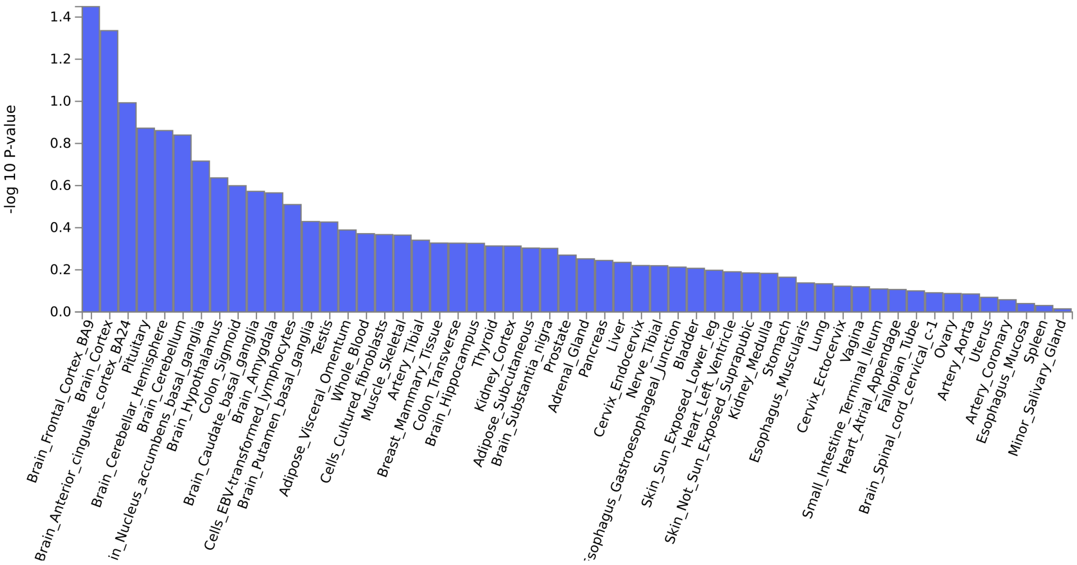


(b) Drinks Per Week WGWAS

Figure S8: Gene tissue expression analysis estimated through MAGMA (implemented in FUMA) using GWAS/WGWAS results for Drinks per week

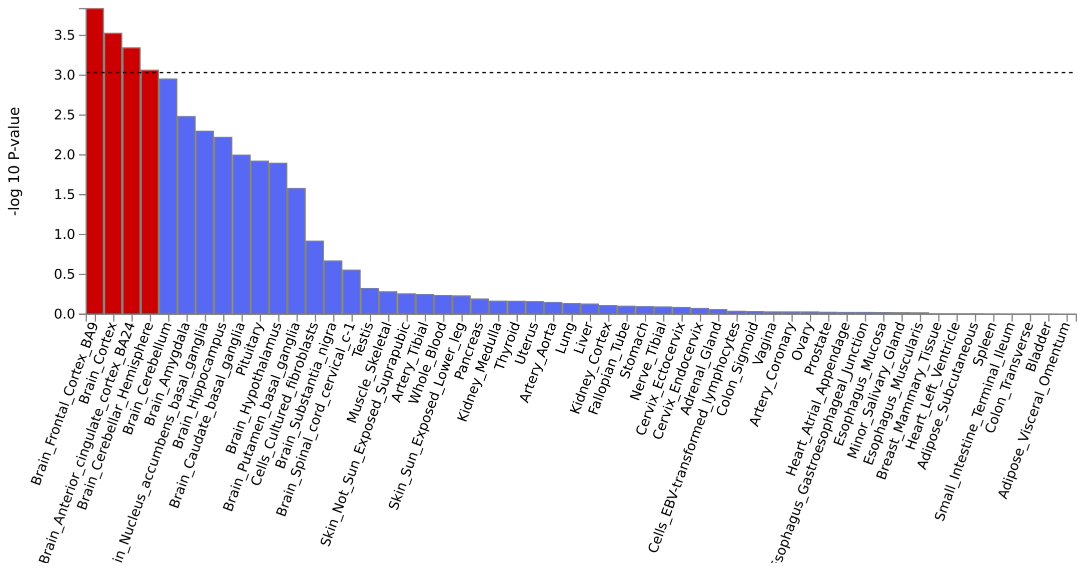


(a) Self-reported health GWAS

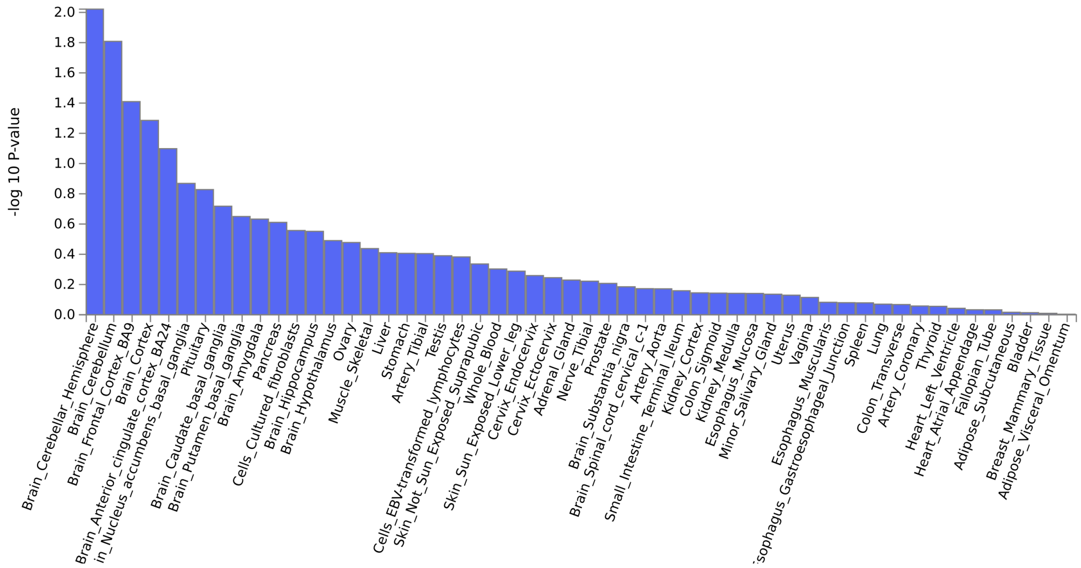


(b) Self-reported health WGWAS

Figure S9: Gene tissue expression analysis estimated through MAGMA (implemented in FUMA) using GWAS/WGWAS results for Self-rated health

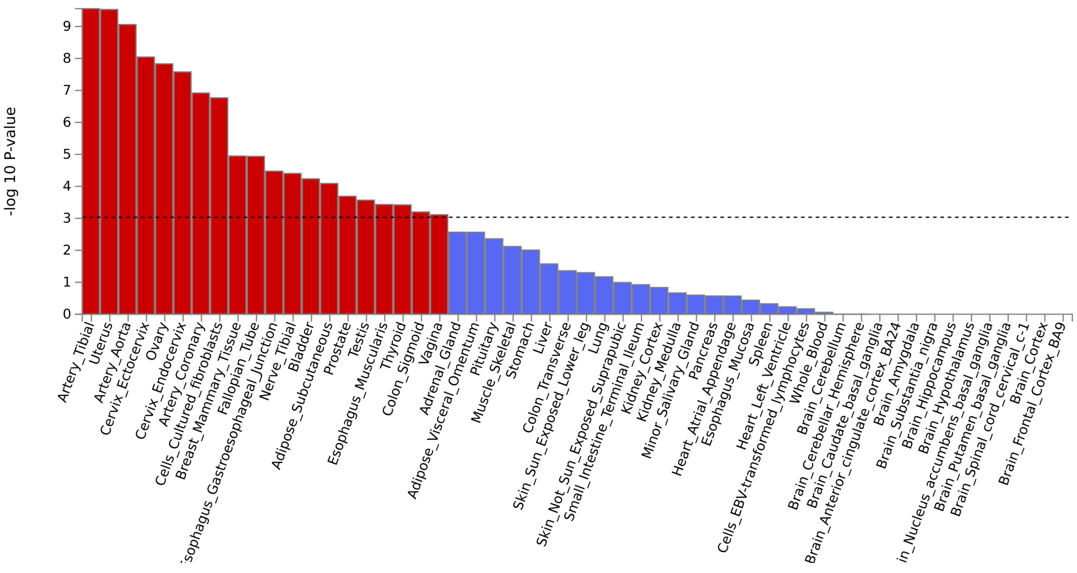


(a) Height GWAS

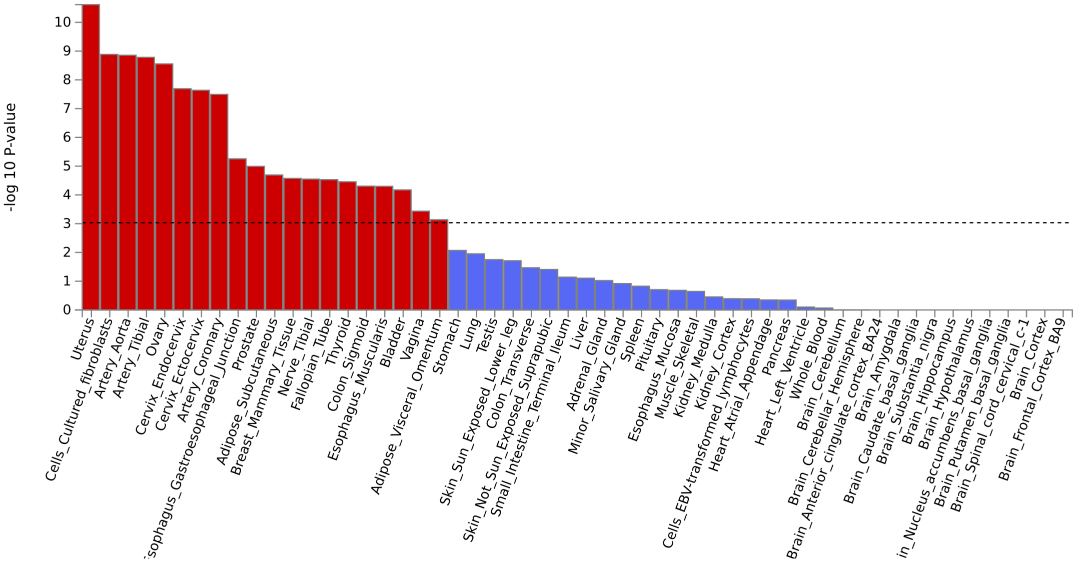


(b) Height WGWAS

Figure S10: Gene tissue expression analysis estimated through MAGMA (implemented in FUMA) using GWAS/WGWAS results for Height

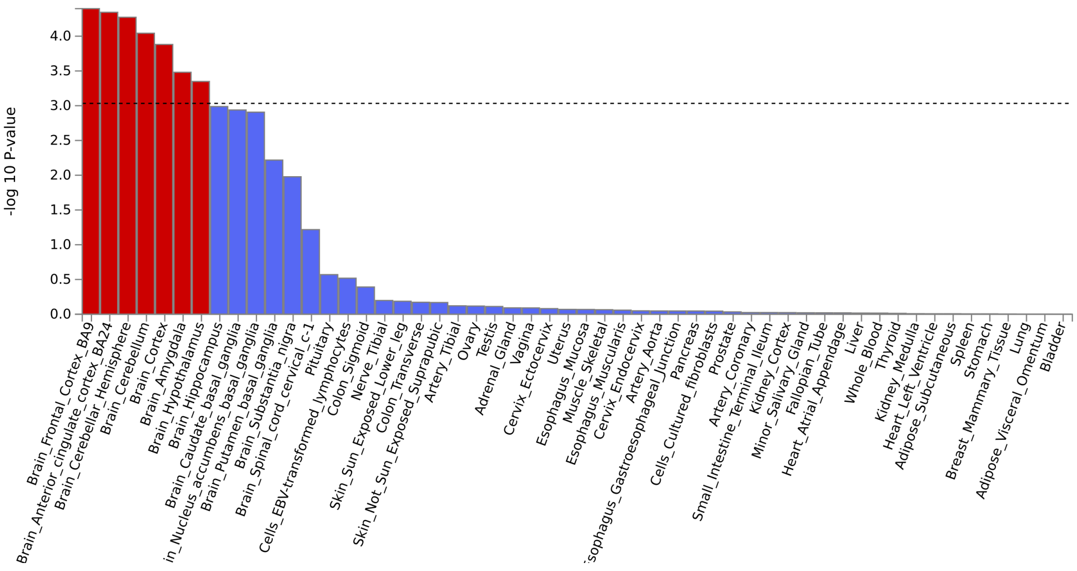


(a) Physical Activity GWAS

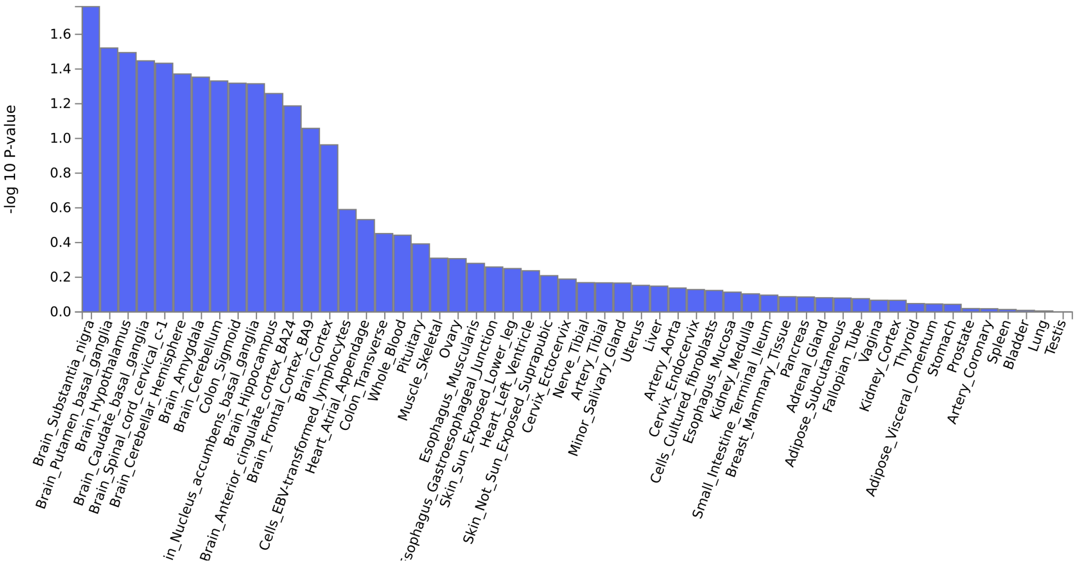


(b) Physical Activity WGWAS

Figure S11: Gene tissue expression analysis estimated through MAGMA (implemented in FUMA) using GWAS/WGWAS results for Physical Activity

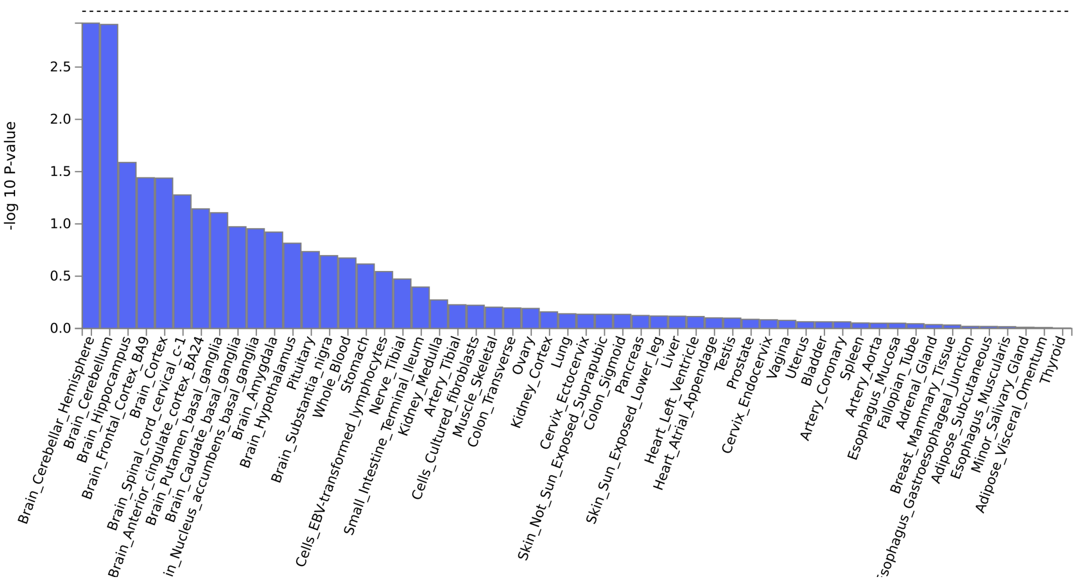


(a) Severe Obesity GWAS

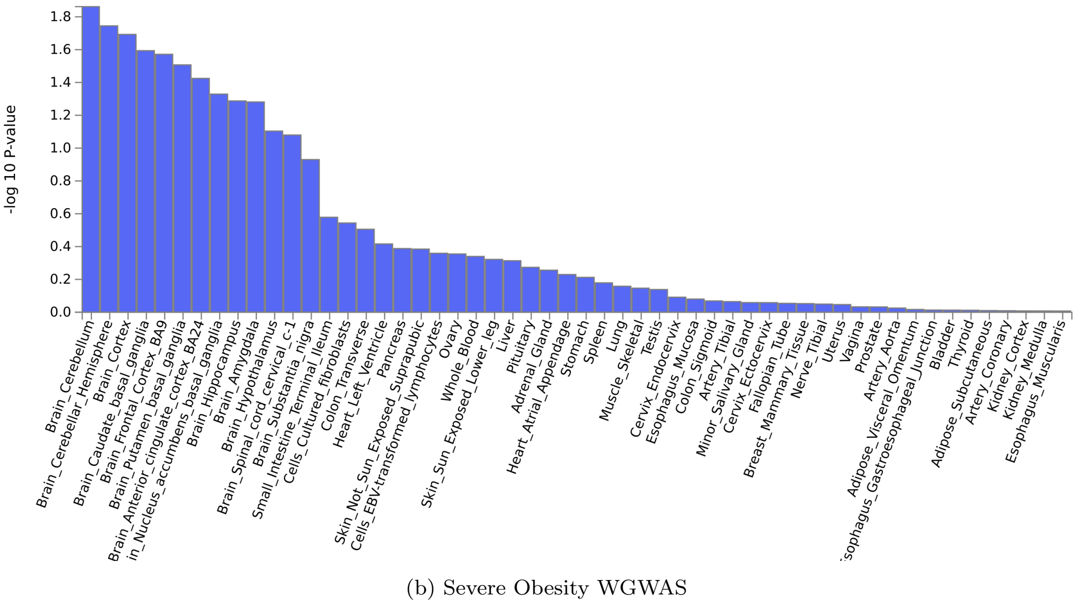


(b) Severe Obesity WGWAS

Figure S12: Gene tissue expression analysis estimated through MAGMA (implemented in FUMA) using GWAS/WGWAS results for Severe obesity

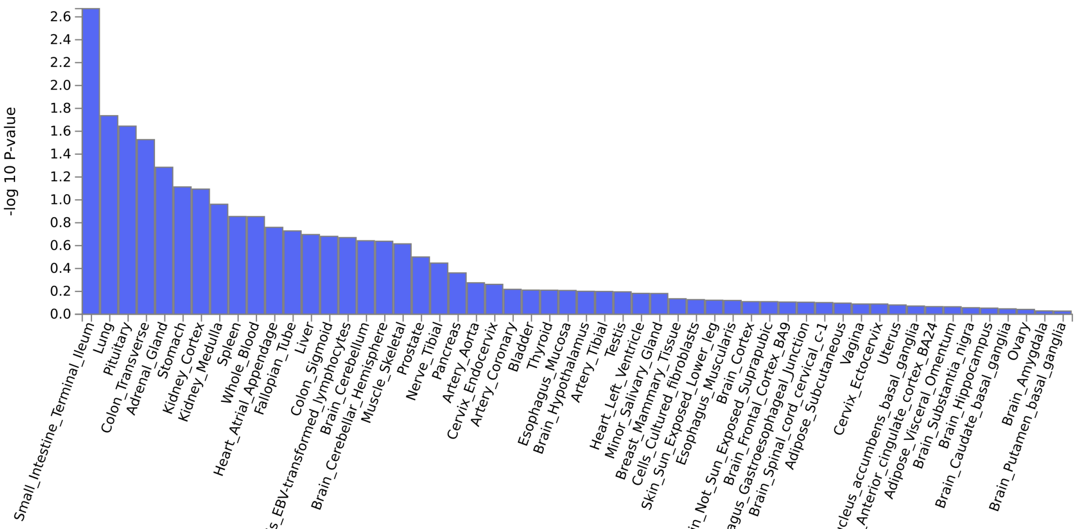


(a) Type 1 Diabetes GWAS

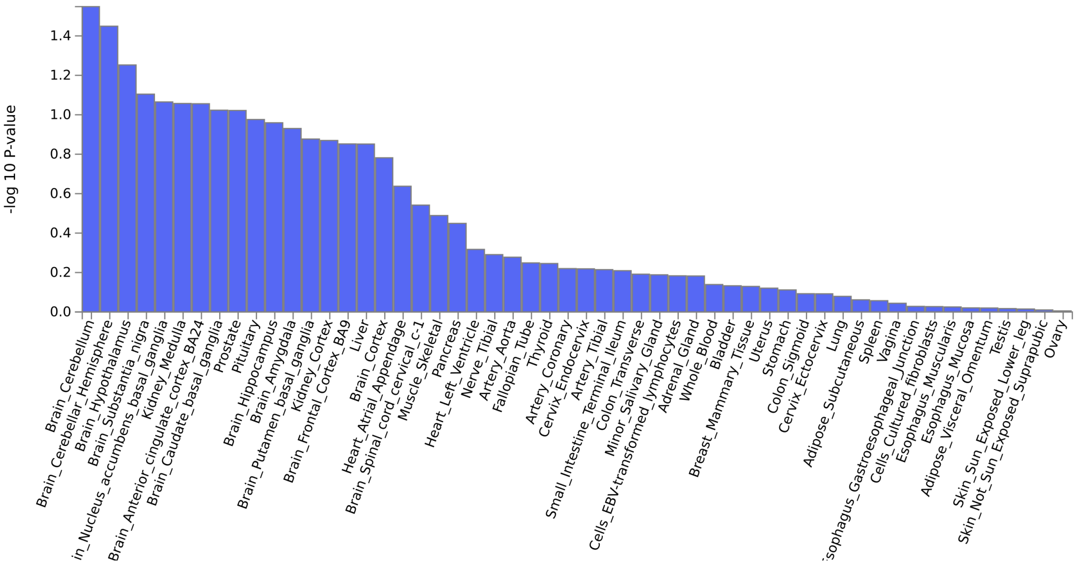


(b) Type 1 Diabetes WGWAS

Figure S13: Gene tissue expression analysis estimated through MAGMA (implemented in FUMA) usingG-WAS/WGWAS results for Type 1 diabetes

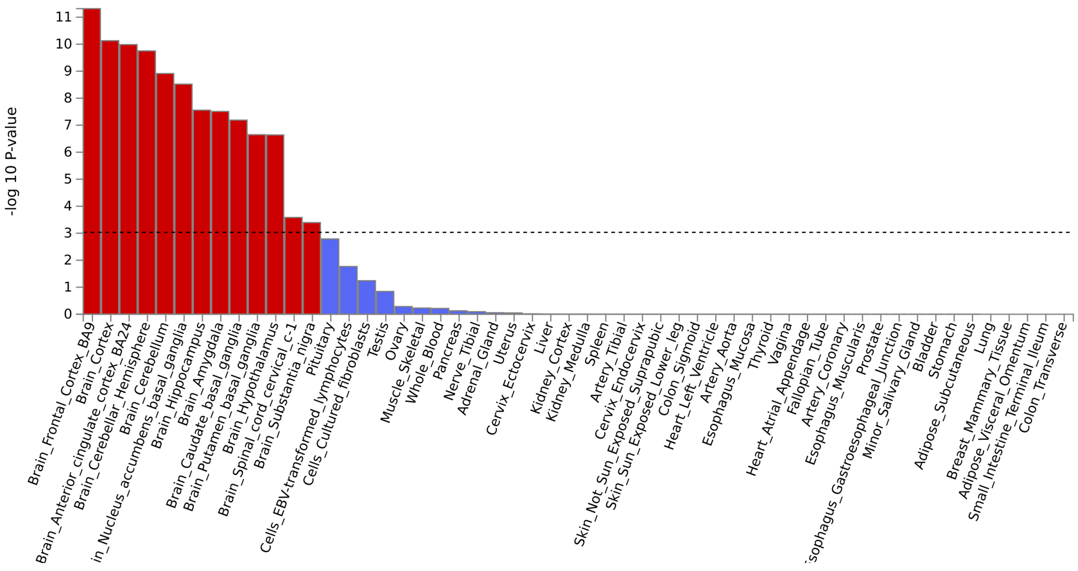


(a) Years of Education GWAS

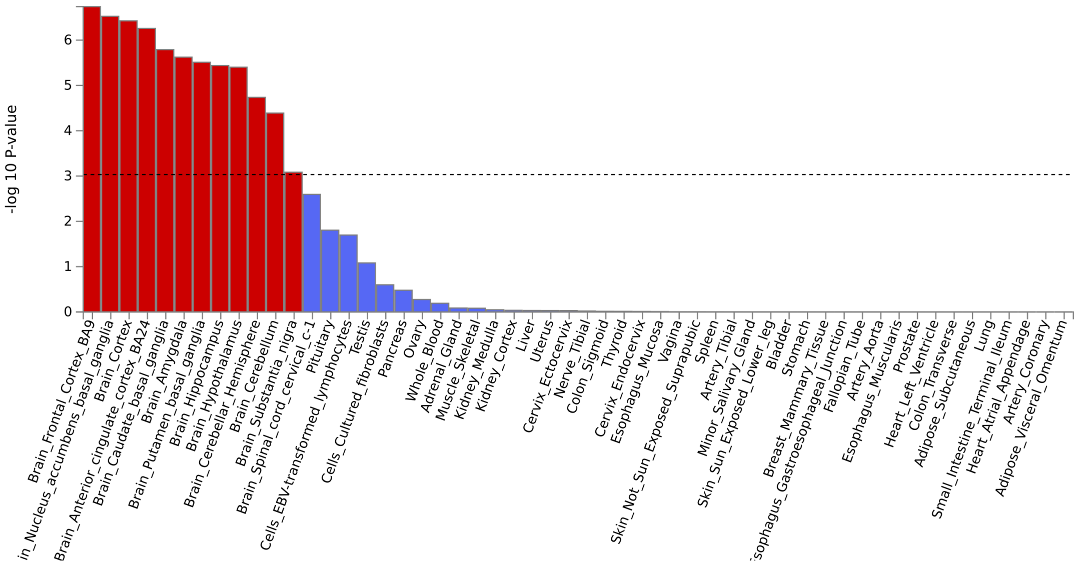


(b) Years of Education WGWAS

Figure S14: Gene tissue expression analysis estimated through MAGMA (implemented in FUMA) using GWAS/WGWAS results for Years of Education

Figure S15: **Zoomed in Manhattan plots of GWAS associations with the UKB inverse probability weights.** Here, we focus on *all* SNPs that are in linkage disequilibrium ($R^2 > 0.1$, 500 kb) with one of the 7 identified lead SNPs for the IP weights ($P < 5 \cdot 10^{-8}$). SNPs that have been found to significantly associate with other traits as found in the GWAS catalog ($P < 5 \cdot 10^{-8}$) are annotated with this trait. The dotted horizontal line shows the genomewide significance level on the negative log scale. Each dot in the plot shows the p-value and base pair position of the association between a SNP (in linkage disequilibrium with the lead SNP) and the IP weights. The lead SNP is depicted as the cross. Each dot is colored by the level of linkage disequilibrium with this lead SNP, as measured by the $R^2$.

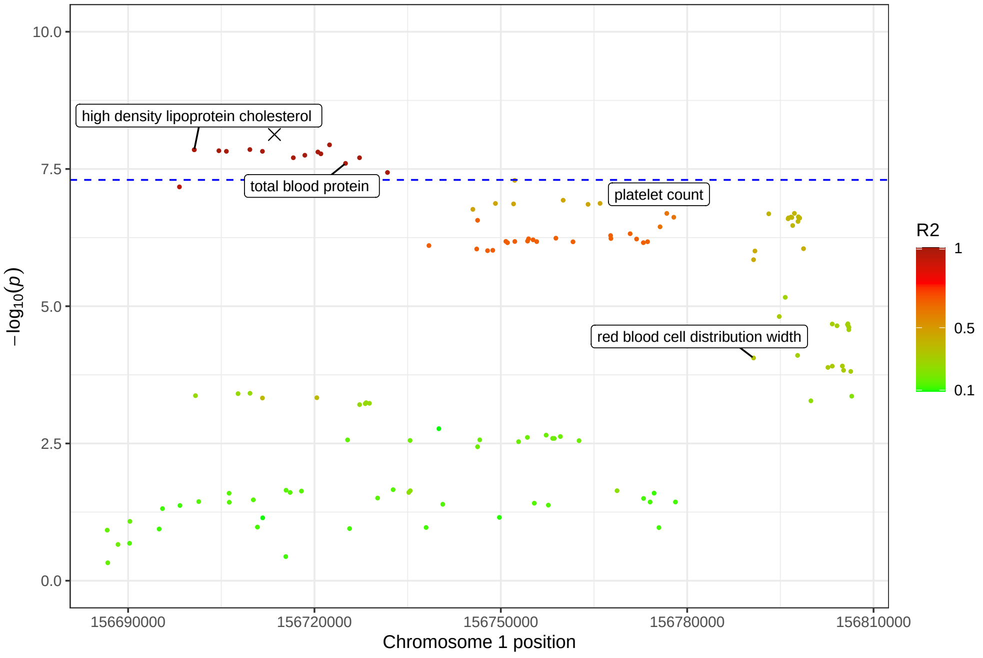


(a) lead SNP rs4399146

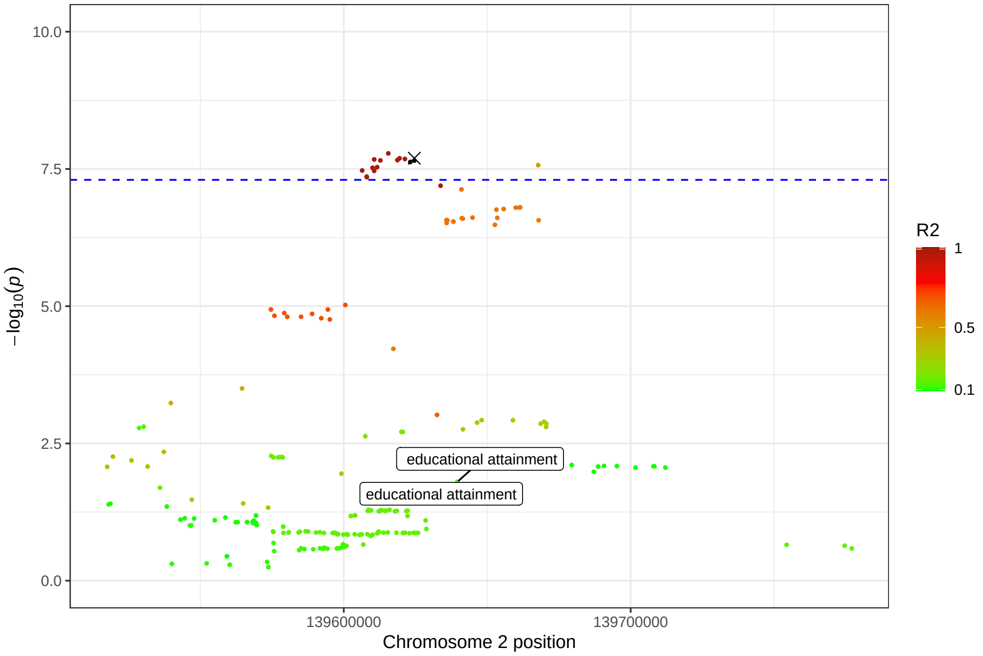


(b) lead SNP rs11885104

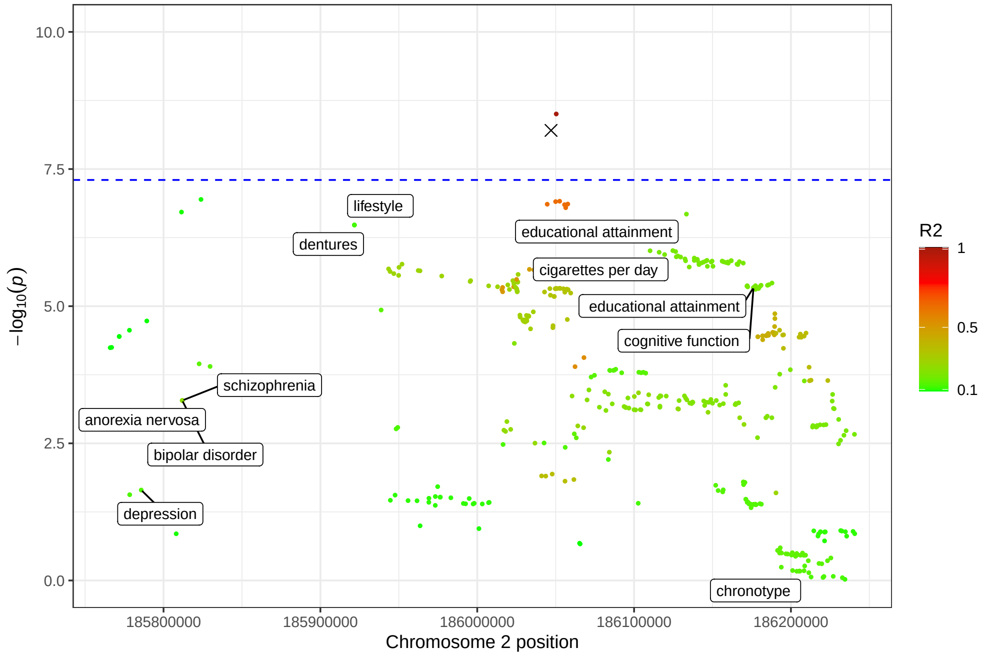


(c) lead SNP rs1483245

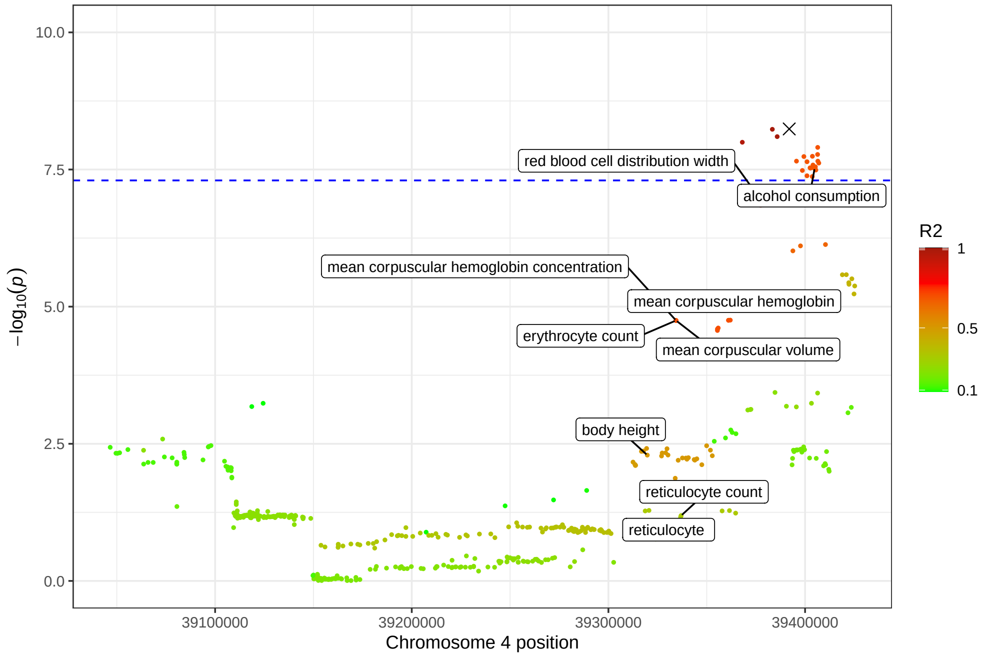


(d) lead SNP rs10033019

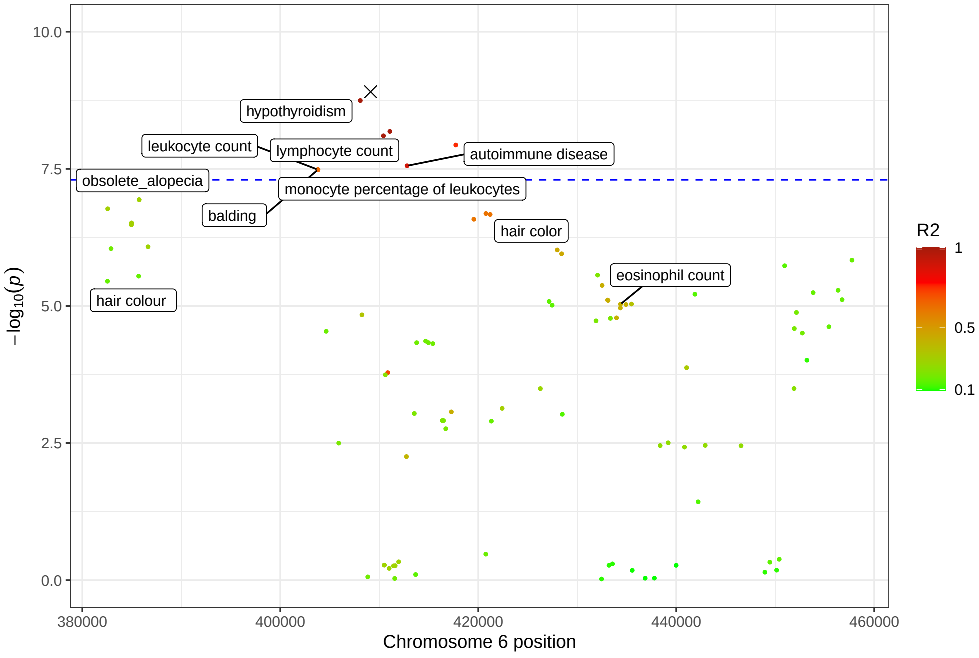


(e) lead SNP rs9391997

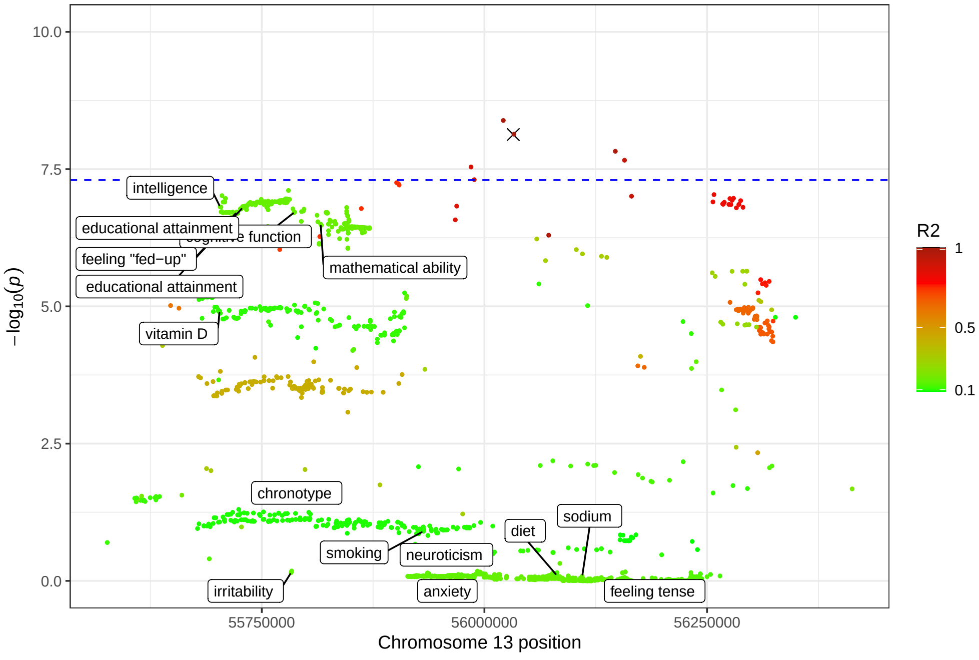


(f) lead SNP rs3013342

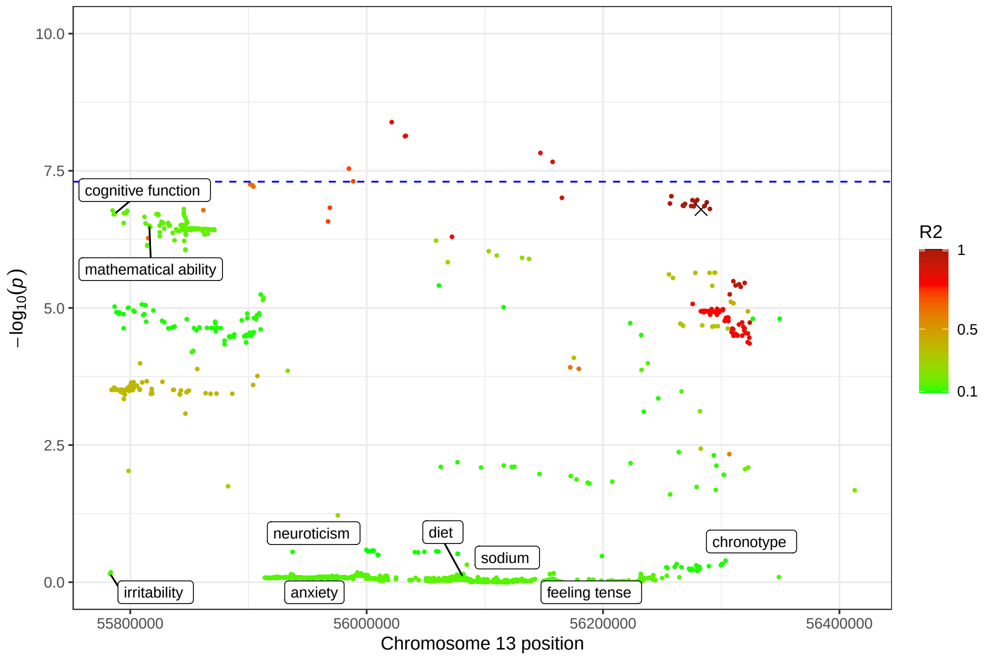


(g) lead SNP rs9597244

Figure S16: **Zoomed in Manhattan plots of WGWAS associations with type 1 diabetes.** Here, we focus on *all* SNPs that are in linkage disequilibrium ($R^2 > 0.1$, 500 kb) with one of the 3 newly identified lead SNPs for type 1 diabetes as found in WGWAS ($P < 5 \cdot 10^{-8}$ in WGWAS *and* $P_H < 5 \cdot 10^{-8}$). SNPs that have been found to significantly associate with other traits as found in the GWAS catalog ($P < 5 \cdot 10^{-8}$) are annotated with this trait. The dotted horizontal line shows the genomewide significance level on the negative log scale. Each dot in the plot shows the p-value and basepair position of the association between a SNP (in linkage disequilibrium with the lead SNP) and type 1 diabetes. The lead SNP is depicted as the cross. Each dot is colored by the level of linkage disequilibrium with this lead SNP, as measured by the $R^2$.

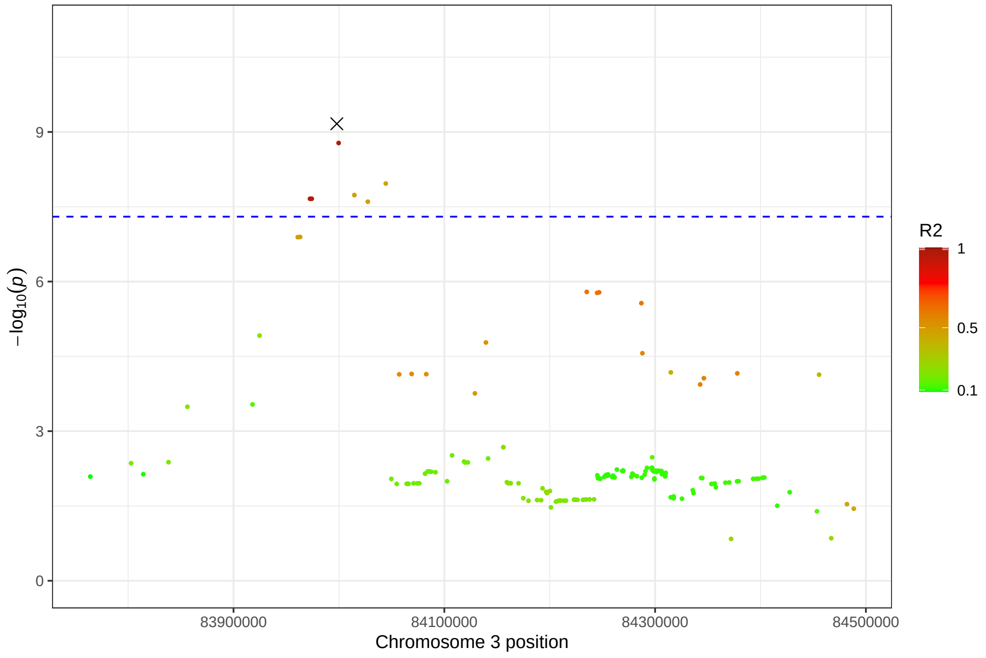


(a) lead SNP rs9861858

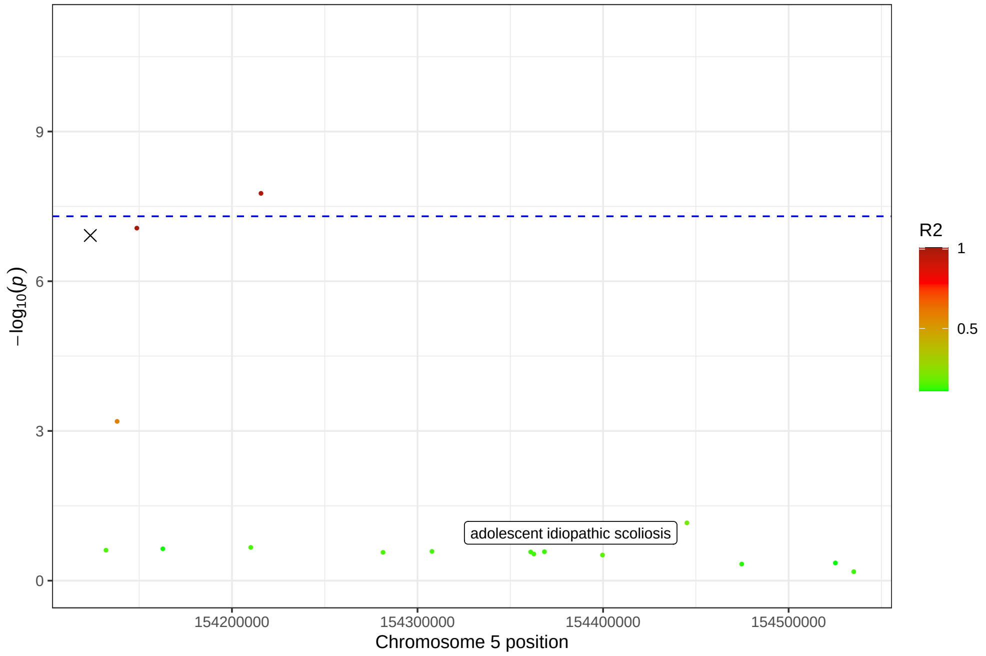


(b) lead SNP rs12522568

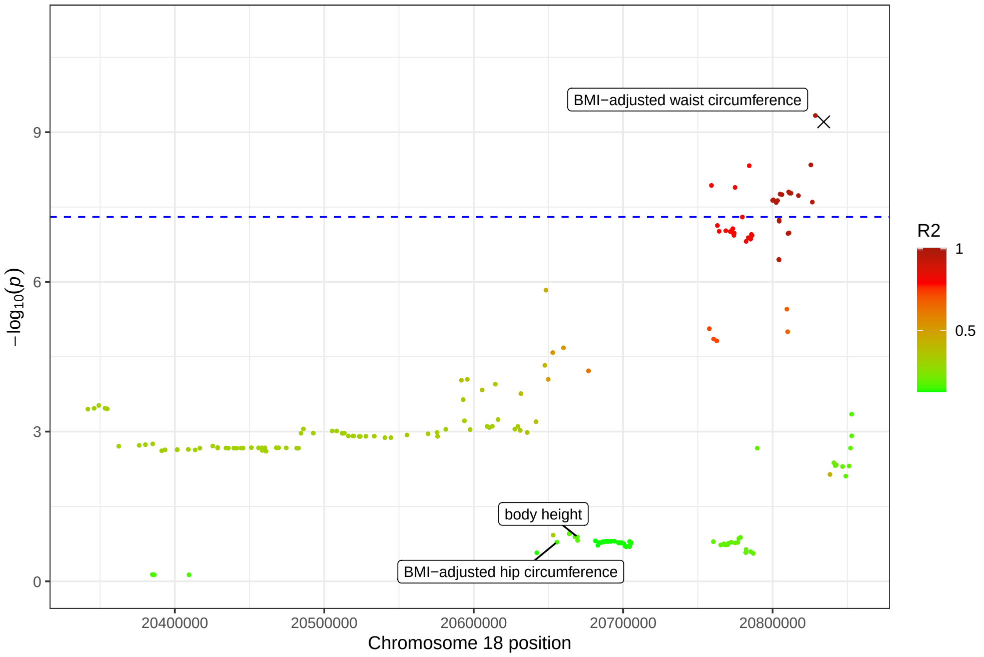


(c) lead SNP rs17186868

Figure S17: **Zoomed in Manhattan plot of WGWAS associations with breast cancer.** Here, we focus on *all* SNPs that are in linkage disequilibrium ($R^2 > 0.1$, 500 kb) with the newly identified lead SNP rs2306412 as found in WGWAS ($P < 5 \cdot 10^{-8}$ in WGWAS *and* $P_H < 5 \cdot 10^{-8}$). None of these SNPs were found to significantly associate with other traits as found in the GWAS catalog ($P < 5 \cdot 10^{-8}$). The lead SNP is depicted as the cross. Each dot is colored by the level of linkage disequilibrium with this lead SNP, as measured by the $R^2$.

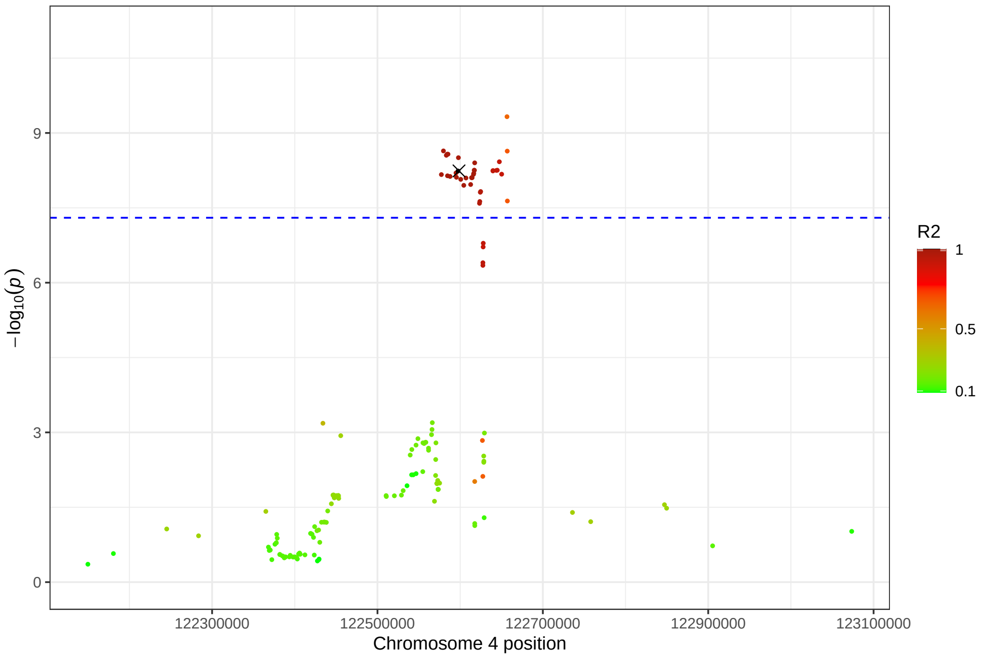

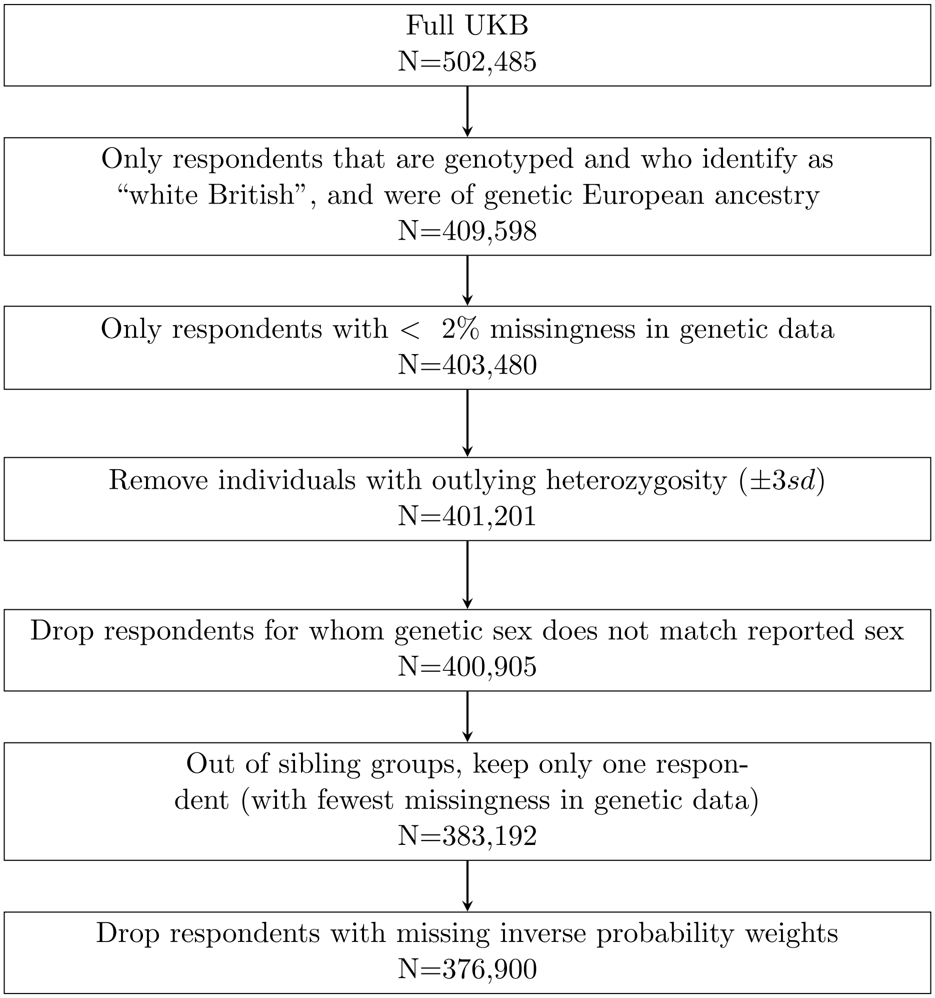


Figure S18: **Summary of sample restrictions made to the UKB**
